# Systematic comparison of family history and polygenic risk across 24 common diseases

**DOI:** 10.1101/2022.07.06.22277333

**Authors:** Nina Mars, Joni V. Lindbohm, Pietro della Briotta Parolo, Elisabeth Widén, Jaakko Kaprio, Aarno Palotie, FinnGen, Samuli Ripatti

## Abstract

Family history is the standard indirect measure of inherited susceptibility in clinical care, while polygenic risk scores (PRS) have more recently demonstrated potential for more directly capturing genetic risk in many diseases. No studies have systematically compared how these overlap and complement each other across common diseases. Within FinnGen (N=306,418), we leverage family relationships, up to 50 years of nationwide registries, and genome-wide genotyping to examine the interplay of family history and genome-wide PRSs. We explore the dynamic for three types of family history across 24 common diseases: first- and second-degree family history, and parental causes of death. Covering a large proportion of the burden of non-communicable diseases in adults, we show that family history and PRS are independent and not interchangeable measures, but instead provide complementary information of inherited disease susceptibility. The PRSs explained on average 10% of the effect of first-degree family history, and first-degree family history 3% of PRSs, and PRS effects were independent of both early- and late-onset family history. The PRS stratified the risk similarly in individuals with and without family history. In most diseases, including coronary artery disease, glaucoma, and type 2 diabetes, a positive family history with a high PRS was associated with a considerably elevated risk, whereas a low PRS compensated completely for the risk implied by positive family history. This study provides a catalogue of risk estimates for both family history of disease and PRSs, and highlights opportunities for a more comprehensive way of assessing inherited disease risk across common diseases.

## INTRODUCTION

Family history (FH) is a risk factor in most common, non-communicable diseases.^1^ With multiple advantages including low cost and non-invasiveness, it captures both genetic and non-genetic familial risk and is therefore widely applied for risk stratification and health promotion. Common clinical applications include assessment of FH of breast cancer for targeted screening, earlier initiation of cardiovascular disease prevention, and evaluating the likelihood of rheumatic disease in patients with inflammatory arthritis.^2-4^ Despite the advantages, assessment of FH also has important limitations in capturing inherited disease risk. Many individuals with common diseases have no FH, or may not know the diseases their relatives have, and the same level of familial risk is assigned to all relatives of similar degree. The accuracy of FH is fairly low owing to factors such as recall bias, and sensitivity to wording in queries may lead to misinterpretation of risk.^5,6^ With average family sizes declining in many developed countries,^7^ FH will also provide increasingly less information for a comprehensive assessment of familial risk.

The algorithmic developments and rapid growth in genome-wide genetic testing provides a more personalized approach for measuring genetic susceptibility through polygenic risk scores (PRS).^8,9^ PRSs employ information from large-scale genetic screens comparing allele frequencies in thousands of disease cases and healthy controls and have identified numerous genetic loci for virtually all common diseases.^10^ To estimate polygenic risks, the common genetic variation and the effects on the disease risks are integrated into a single metric, the PRS. The effectiveness of PRSs in risk stratification has been demonstrated for many diseases, with predictive value demonstrated alongside established clinical risk assessment tools.^11^ Similarly, PRSs modify risk among carriers of high-risk variants and identify high-risk individuals for whom existing prediction tools are suboptimal.^11-16^

Given the initial expense of implementing PRS estimation in a clinical setting relative to the seemingly simple questions pertaining to family history, systematic evaluation of the independent added benefit of PRS across common diseases is essential. Studies on individual diseases have observed fairly independent effects of PRS and first-degree FH, ^11,15,17-27^ but no studies have systematically compared the relative contributions and overlap of PRS and FH across different types of familial risk, across varying genetic architectures, and across a wide range of diseases. Moreover, only few studies have used genome-wide PRSs, although these contemporary PRSs containing a large number of variants have demonstrated improved performance beyond PRSs with less variants due to high polygenicity in common diseases.^13,28-30^ Here we study the interplay of first and second-degree FH, parental causes of death, and genome-wide PRSs for 24 diseases using FinnGen (N=306,418), showing that FH and PRSs are largely independent and provide complementary information in risk assessment.

## MATERIAL AND METHODS

### Participants and diseases

This observational study uses FinnGen study Data Freeze 7, a collection of 306,418 adults (age ≥18) from epidemiological cohorts, disease-based cohorts, and hospital biobanks (**Supplementary Table 1**). We used three binary definitions for FH: 1) any type of first-degree FH (FH_1st_ morbidity or mortality), 2) any type of second-degree FH (FH_2nd_), and 3) parental cause of death (FH_P_). Both for the index patients and their relatives (i.e. how FH was obtained), cases were identified through nationwide healthcare registries. The 24 diseases were chosen based on availability of large published genome-wide association studies (GWAS) with full summary statistics available for genome-wide PRSs (**Supplementary Table 2)**. Case definitions are in **Supplementary Table 3**. Registry follow-up ended at Dec 31, 2019, with parental causes of death available until Dec 31, 2018. For FH_P_, we studied 15 out of the 24 diseases, identifying causes of death (immediate, contributing, and underlying causes of death). Details on genotypes, PRS generation and inference of relatedness is in Supplementary Methods.

### Polygenic risk scores

For each of the 24 diseases, we constructed disease-specific PRSs in a systematic manner (**Supplementary Table 2)**. The number of cases in the GWASs ranged from 3,769 (epilepsy) to 567,460 (eGFR used for chronic kidney disease). PRS-CS was used for inferring posterior effect sizes for calculation of genome-wide PRSs^31^ with 1000 Genomes Project European sample (N = 503) as the external linkage disequilibrium (LD) reference panel.^32^ The PRS was analyzed primarily as a continuous variable, with selected analyses applying either a 1) binary definition of FH, with high PRS defined as a PRS in the top decile of the distribution, with the rest as the reference group, or 2) PRS categories 0-10%, 10-20%, 20-40%, 40-60%, 60-80%, 80-90%, and 90-100%, with the reference group being 40-60%. To assess the impact of high vs low PRS, the reference category was 33^rd^ to 90^th^ percentiles, and low PRS was defined as the lowest tertile of the distribution, to allow for a sufficient number of cases with low PRS.

### Statistical analysis

Associations between FH, PRS, and risk of disease was assessed with logistic regression, with models adjusted for sex, birth year, genotyping array, cohort, and the first ten genetic principal components of ancestry. The same adjustments were applied when calculating area under the receiver operator characteristic curve (AUC) estimates. AUC estimates with 95% CIs were calculated with R package pROC. Interactions between FH and the continuous PRS (scaled to zero mean and unit variance) was assessed by introducing their interaction term to the regression model, assessing statistical significance set at a p-value threshold of 0.0013 (Bonferroni-correction for 24+15 tests). Cumulative incidences by age 80 were estimated with Kaplan-Meier survival curves (R package survminer). Statistical analyses were performed using R, version 4.1.0.

## RESULTS

FinnGen comprises 306,418 individuals (56.3% women; mean age 59.8 at the end of follow-up in 2019, s.d. 17.3). For the 24 diseases, FH was defined as 1) first-degree family history, FH_1st_ (morbidity or mortality), 2) second-degree family history, FH_2nd_, and 3) parental cause of death, FH_P_. Each identifies the relatives’ diagnoses systematically through nationwide registries, including the hospital discharge registry (available from 1968-), causes of death registry (1964-), the Finnish Cancer Registry (1953-). FH_1st_ and FH_2nd_ leverage the genetic relatedness within FinnGen: out of 306,418 individuals, we identified 39,444 with first-degree relative pairs based on the KING kinship coefficient^33^ (see methods for details; 60.3% women; mean age 53.0, s.d. 16.5; parent-offspring relationship in 19,261 individuals, full-sibling in 20,183). For breast cancer, we studied only women (15,281 individuals, mother-daughter relationship in 7,770; full sisters in 7,511) and for prostate cancer, only men (9,473 individuals; father-son relationship in 3,932; full brothers in 5,541). Similarly, we identified 47,154 individuals with a second-degree relative in the dataset (63.2% women, mean age 47.5, s.d. 15.0; N=18,973 for breast cancer; N=12,355 for prostate cancer). Parental causes of death (FH_P_), were linked through causes of death registry available from 1964 to 2019 and we excluded 78,436 whose both parents have died before 1964 or who had missing data on both parents (e.g. due to emigration), resulting in 227,982 individuals (mean age 53.6, s.d. 15.1; N=133,653 for breast cancer; N=94,329 for prostate cancer; 70,225 (30.1%) with one and 73,299 (32.2%) with two dead parents). See **Supplementary Figure 1** for study flow diagram.

### Family history and risk of disease

First, we systematically evaluated the effects of FH on risk of disease. **Figure 1** shows the prevalence of the diseases, and the prevalence and effect sizes for positive FH. The most common diseases were cardiometabolic diseases, followed by knee osteoarthritis and hypothyroidism. Positive FH_1st_ was significantly associated with higher risk of disease in all 24 diseases except stroke. The effect sizes ranged from odds ratio (OR) 3.25 (95% confidence interval, CI, 2.41–4.37) in chronic kidney disease to OR 1.17 (0.98–1.39) in stroke (**Supplementary Table 4**). For FH_2nd_, 18/24 diseases showed evidence of an association, with their effect sizes ranging from OR 1.85 (1.19–2.89) in colorectal cancer to OR 1.17 (1.09–1.25) in hypertension (**Supplementary Table 5)**. Compared to FH_1st_, the effect sizes for FH_2nd_ were on average 69.1% lower (s.d. 25.0%; calculated from log odds), i.e. a third of the effect of FH_1st_. For FH_P_, out of the 24 diseases, we studied 15 diseases that are well captured by causes of death and used information from all recorded causes of death (immediate, contributing, and underlying causes of death on the death certificate). For all 15 diseases, we observed an association between FH_P_ and risk of disease, with effect sizes ranging from OR 2.82 (2.25–3.53) in seropositive rheumatoid arthritis to OR 1.12 (1.04-1.20) in stroke (**Supplementary Table 6**). Compared to FH_1st_, the effect sizes for FH_P_ were on average 30.1% lower (s.d. 22.4%), i.e. two thirds of the effect of FH_1st_.

**Figure 1.**
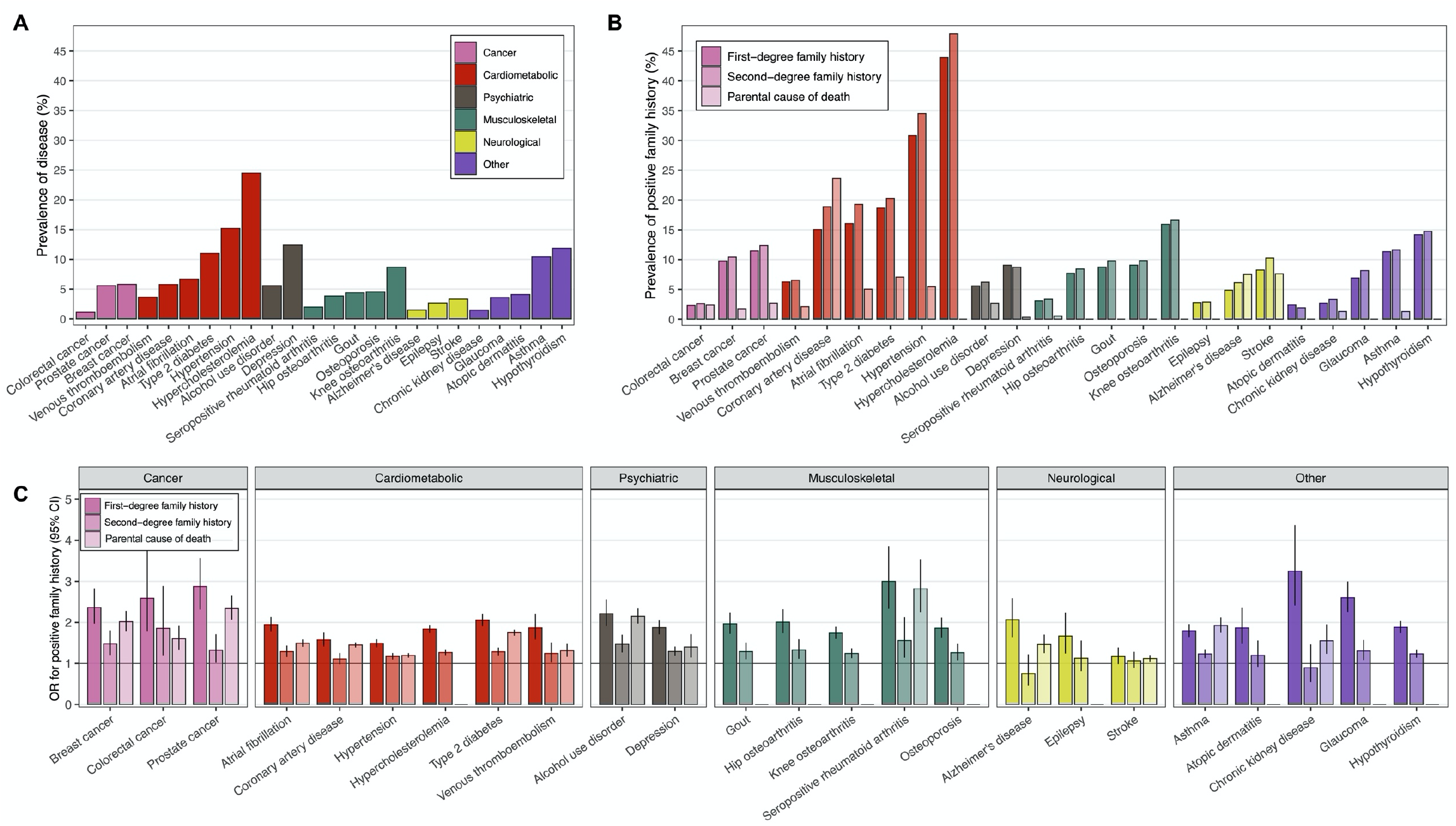
Disease prevalence and prevalence and effect sizes of positive family history. **Panel A:** disease prevalence in individuals for whom we studied risk of first-degree family history **Panel B:** Prevalence of first-degree family history (left column), second-degree family history (middle column), and parental cause of death (right column). **Panel C:** Effect size of first-degree family history (left column), second-degree family history (middle column), and parental cause of death (right column) with respective diseases. For parental causes of death, we studied 15 out of the 24 diseases. Sample size in panel A: total N = 39,444, N = 15,281 for breast cancer, N = 9,473 for prostate cancer. Sample sizes in panels B-C: first-degree family history as in panel A, second-degree family history total N = 47,154, N = 18,973 for breast cancer, N = 12,355 for prostate cancer, and parental causes total N = 227,982, N = 133,653 for breast cancer; N = 94,329 for prostate cancer. Odds ratios (OR) were obtained from logistic regression models adjusted for sex (except for breast and prostate cancer), birth year, genotyping array, cohort, and the first ten genetic principal components of ancestry.

### Overlap of family history and polygenic risk

Next, we compared the overlap between FH and PRSs. We constructed 24 genome-wide PRSs with uniform methodology using PRS-CS^31^, one for each disease (**Supplementary Table 2**). We first compared the effects sizes per standard deviation (SD) increase for PRS, and FH_1st_ (**Figure 2, Supplementary Table 4**). The PRS was associated with elevated risk in all 24 diseases. The higher the PRS, the higher was the proportion of positive FH (**Supplementary Figure 2)**. Effect sizes for the PRS ranged from OR 2.33 (95%CI 2.10–2.558) in prostate cancer to OR 1.12 (1.05-1.20) in epilepsy. Adjusting the PRS effect size with FH_1st_, the change in effect size was small (mean decrease as log odds -3.0%, s.d. 1.3%). Adjusting the effect of FH_1st_ with PRS led to a mean decrease of - 10.3% (s.d. 6.0%), i.e. PRS explained of tenth of first-degree family history. No effect size decrease was observed for PRS adjusting with FH_2nd_ (**Supplementary Table 5**). We observed similar results for FH_P_ (**Supplementary Table 6**; effect size decrease adjusting PRS effects with FH_P_ -0.7%, s.d. 0.6%, vice versa -14.5%, s.d. 9.2%). Proportional decreases in log odds by disease for all definitions of FH are in **Figure 3**. FH generally explained a much smaller fraction of the effect of PRS than vice versa. A similar pattern was observed categorizing the PRS and comparing high PRS (>90^th^ percentile) to the rest of the distribution (**Supplementary Table 7, Supplementary Figure 3**). A high PRS conferred on average similar effect sizes as FH_1st_. The effect sizes particularly in common cancers and cardiometabolic diseases were higher for the PRS, whereas the effect sizes for psychiatric diseases were higher for FH_1st_.

**Figure 2.**
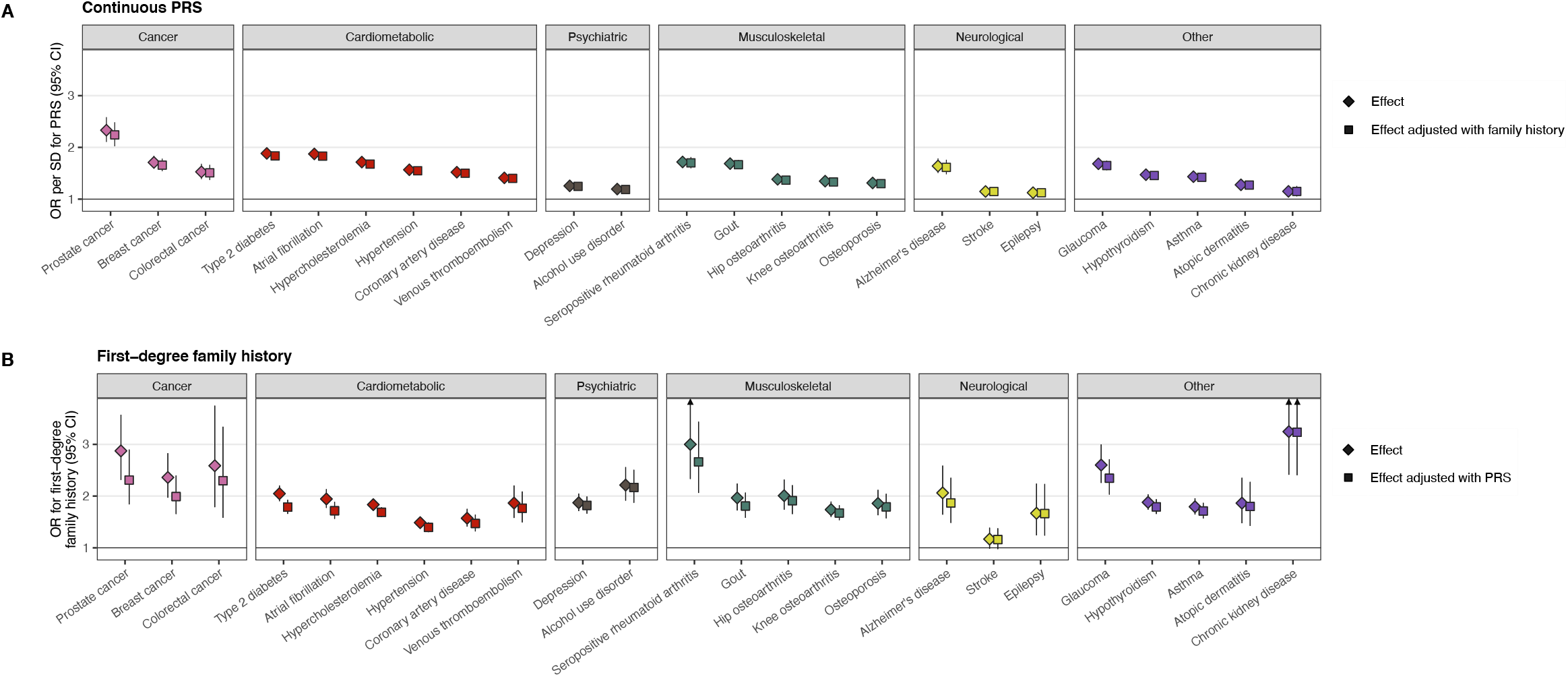
Cross-adjustment effects for first-degree family history (FH_1st_), and respective polygenic risk scores (PRS). The impact of adjusting the PRS effect with first-degree FH_1st_ **(panel A)** and vice versa **(panel B)**. The diamonds represent the unadjusted effects and the squares the adjusted effects. The PRS explained on average 10% of the effect of FH_1st_, but FH_1st_ only 3% of the PRSs. The PRS effect is shown per one SD increase. Total N = 39,444, N = 15,281 for breast cancer, N = 9,473 for prostate cancer. Odds ratios (OR) were obtained from logistic regression models adjusted for sex (except for breast and prostate cancer), birth year, genotyping array, cohort, and the first ten genetic principal components of ancestry.

**Figure 3.**
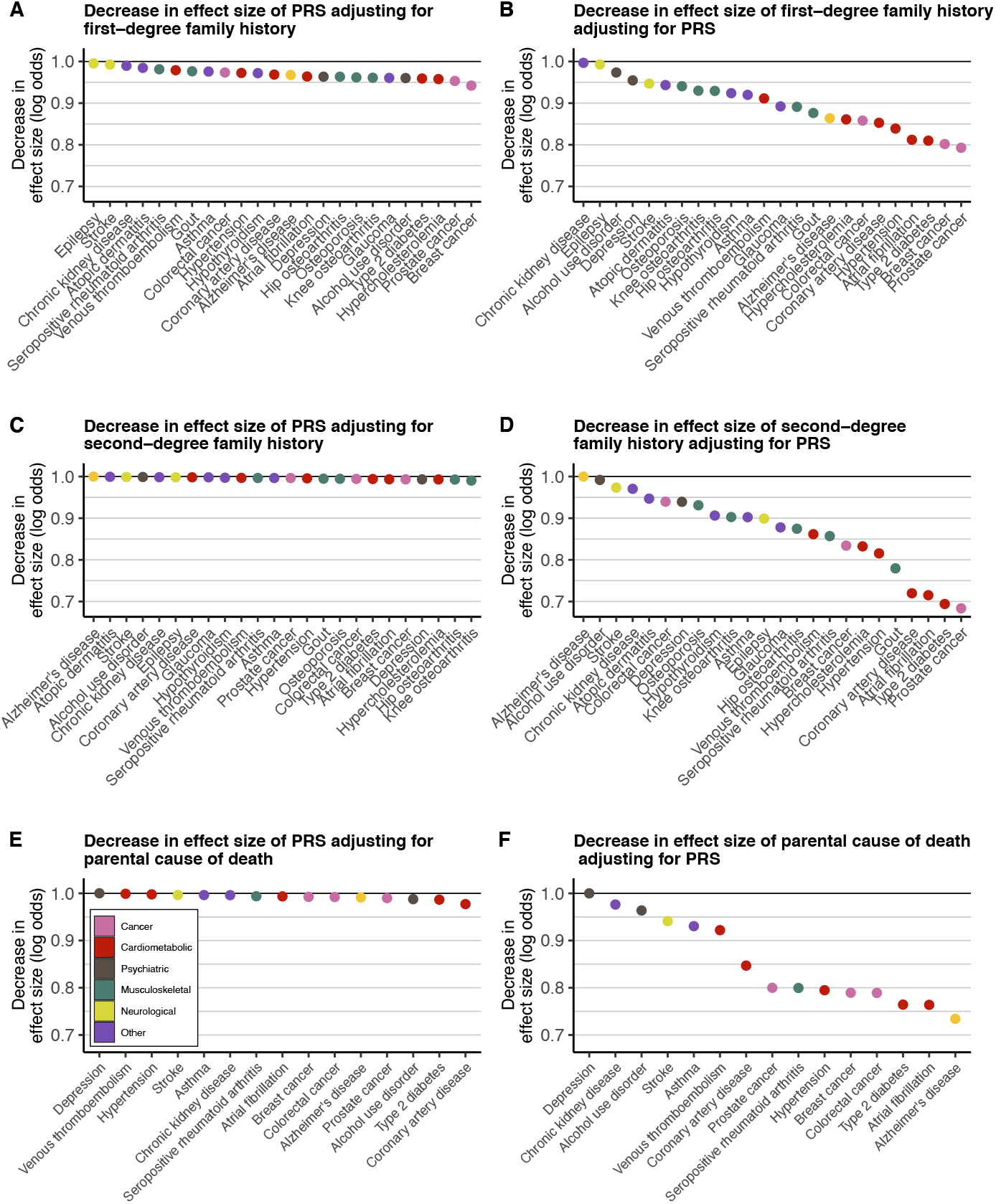
Proportional decreases in log odds by disease for first-degree family history FH_1st_ (panels **A-B**), for second-degree family history FH_2nd_ (panels **C-D**), and parental causes of death FH_P_ (panels **E-F**). The left column (panels **A, C, E**) represents decreases in effect size of high polygenic risk score (PRS; defined as top 10% of the distribution) adjusting for family history. The right column (panels **B, D, F**) represents decreases in effect size of family history adjusting for high PRS. The y axis represents the decrease in the effect size, calculated by dividing the log odds from the adjusted logistic regression model with the log odds from the non-adjusted model. For instance, in panel **A**, the y axis represents the following quantity: (log odds of PRS adjusting for FH_1st_) / (log odds of PRS without adjusting for FH_1st_). The reference category for PRS was the rest of the distribution (individuals below the 90^th^ percentile). In panel D, the proportion of Alzheimer’s disease was set at 1.00 as we did not observe any association for second-degree family history of Alzheimer’s disease.

As early-onset FH is considered a particularly important familial risk factor, we also assessed the impact of FH_P_ divided into tertiles of age at death. The largest effect size was observed for FH_P_ with the lowest age tertile, in line with early-onset FH being a stronger risk factor than late-onset FH. Adjusting the PRS with this FH_P_ divided into age tertiles had no impact on the effect sizes of the PRSs. Adjusting this FH_P_ by PRS resulted in the largest effect size decreases for the youngest age tertile, but the decreases were overall small. These show that the PRS was independent of both early- and late-onset FH_P_ **(Supplementary Table 8, Figure 4**).

**Figure 4.**
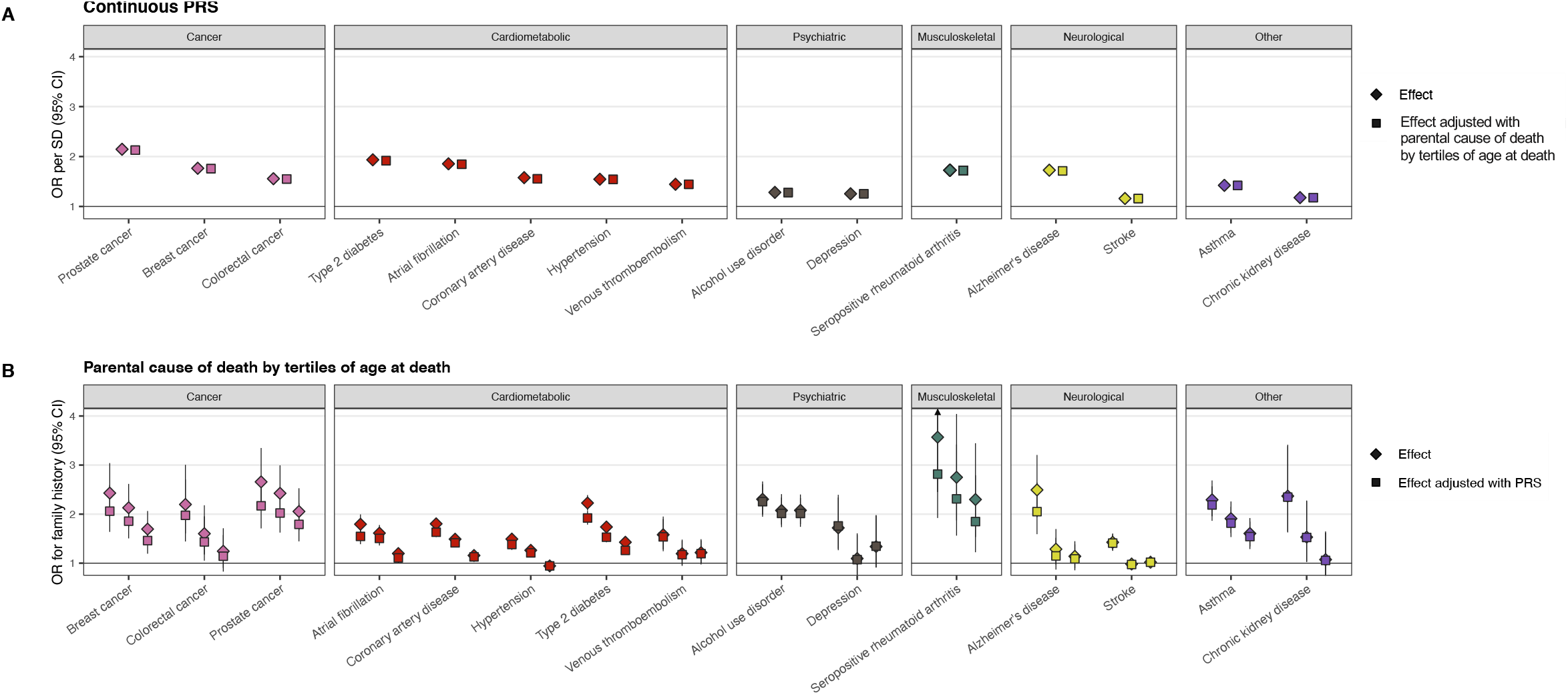
Polygenic risk is independent of both early- and late-onset family history. As early-onset family history is considered a particularly important familial risk factor, we also assessed the impact of FH_P_ divided into tertiles of age at death. **Panel A:** Adjusting the effect of polygenic risk score (PRS; per SD) by parental causes of death (FH_P_) divided into tertiles of age at death had no impact on the effect sizes of the PRSs. **Panel B:** Adjusting the effects FH_P_ by tertiles of age at death by PRS resulted in the largest effect size decreases for the youngest age tertile, however, for most diseases the difference by age tertile was small. The diamonds represent the unadjusted effects and the squares the adjusted effects. In panel B, the effect sizes from with lowest to highest age at death are displayed from left to right, and the reference group for each disease is individuals with negative FH_P_. Sample size: total N = 227,982; N = 133,653 for breast cancer; N = 94,329 for prostate cancer. Odds ratios (OR) were obtained from logistic regression models adjusted for sex (except for breast and prostate cancer), birth year, genotyping array, cohort, and the first ten genetic principal components of ancestry. Age limits for tertiles of FH_P_, and the number of individuals with parental cause of death in each tertile are reported in Supplementary Table 7.

With formal interaction testing, we did not identify any systematic interactions between FH and PRS (**Supplementary Figure 4**), which was further supported by observing similar PRS effect sizes in individuals with positive and negative FH_1st_ (**Figure 5**).

**Figure 5.**
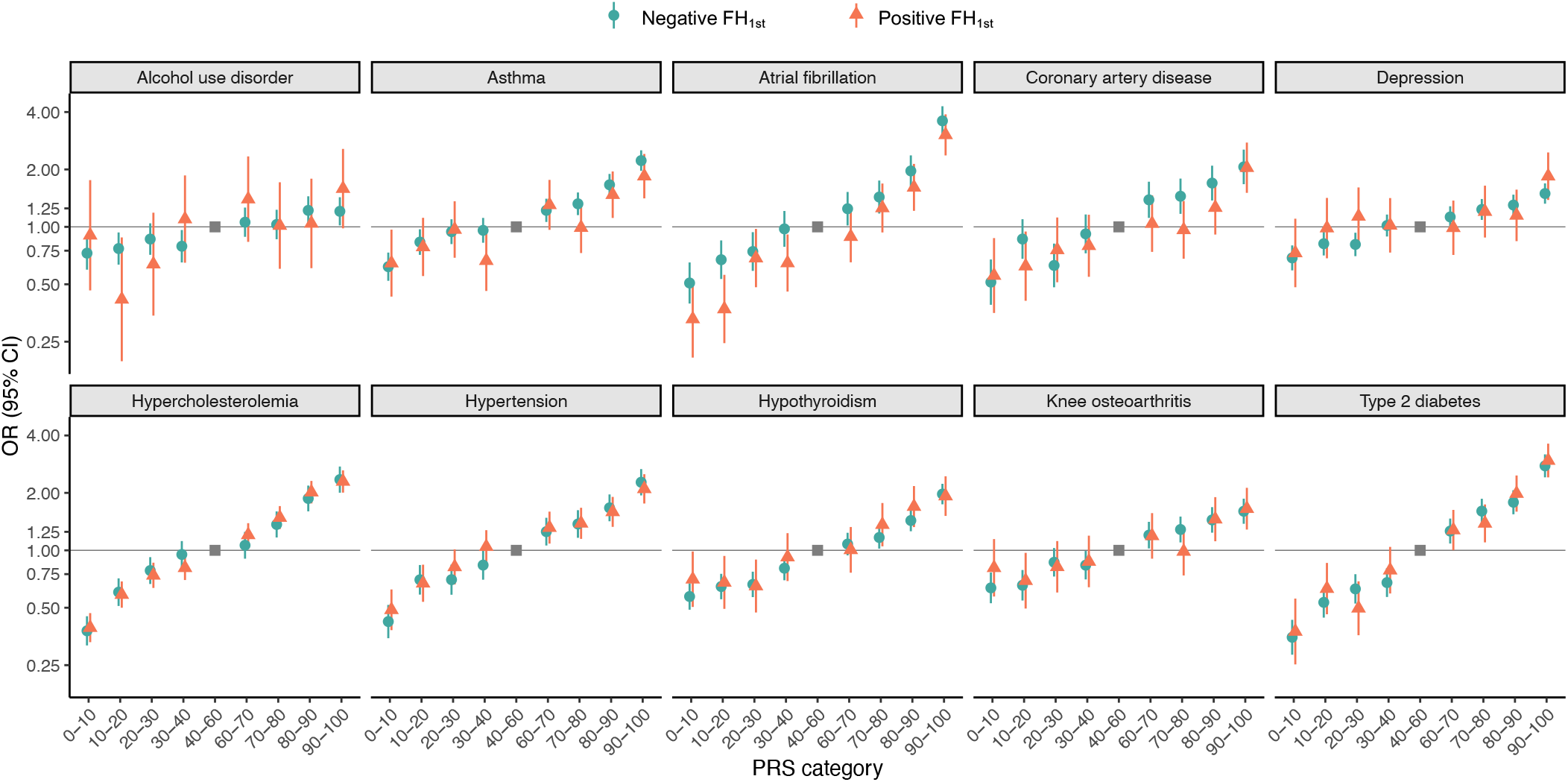
Effect sizes of polygenic risk scores stratified by first-degree family history (FH_1st_), assessed for the 10 most prevalent diseases.

### Polygenic risk in individuals with a positive family history

Next, having assessed the overlap between FH and the PRSs, we estimated how high and low PRS impact disease risk in individuals with positive FH_1st_. Looking at cumulative incidence of risk of disease with the PRS divided into three groups (high PRS >90%, average PRS 33-90%, and low PRS <33%), we observed that a low PRS systematically compensated for the impact of positive FH_1st_, and individuals with a combination of high PRS and positive FH_1st_ had a particularly high risk (**Figure 6**). Survival curves for a broader set of diseases, and survival curves stratifying individuals with no FH_1st_ into similar PRS groups are in **Supplementary Figures 5–6**.

**Figure 6.**
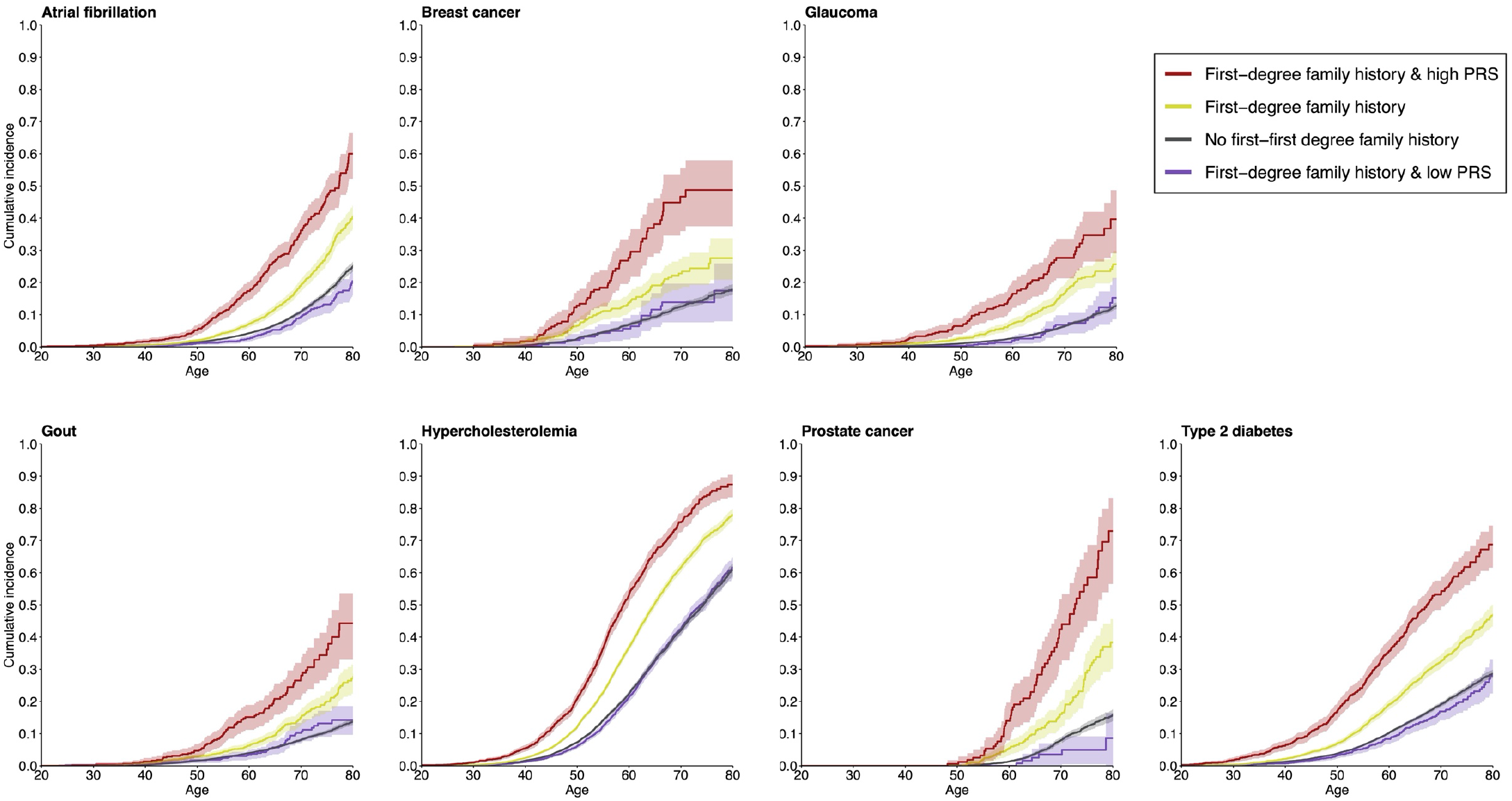
The impact of polygenic risk on disease risk in individuals with positive family history. The survival curves show cumulative incidences for individuals with positive first-degree family (FH_1st_), stratified by level of polygenic risk score (PRS). High PRS was defined as top decile of the PRS distribution and low PRS as the bottom tertile of the PRS distribution. The figure shows results for the five diseases with the largest effect sizes for PRS, and for breast and prostate cancer. Survival curves for a broader set of diseases, and survival curves stratifying individuals with no FH_1st_ into similar PRS groups are in **Supplementary Figures 5–6**. Total N = 39,444, N = 15,281 for breast cancer, N = 9,473 for prostate cancer. Analyses were performed for diseases with an OR >2 for high PRS in Figure 2 (15/24 diseases), and over 10 cases in each subgroup, excluding Alzheimer’s disease due to its average onset late in life.

### Concordance of high polygenic risk in relatives

Lastly, we assessed concordance – detection of a high PRS among first- and second-degree relatives, relevant for cascade screening in relatives of individuals with high PRS. We evaluated two questions: 1) “What is the probability of having high PRS, if a relative has high PRS?” and 2) “How does this probability differ with relative’s disease status?”. For 1), on average 33.7% of the first-degree and 19.8% of second-degree relatives had a similarly high PRS (**Supplementary Figures 7–8**). For 2), the concordance was somewhat higher with positive FH_1st_ than with negative FH_1st_, with an average difference of 2.5% (range 0.0–7.9%). For FH_2nd_, no difference with disease status was observed (average 0.6%).

## DISCUSSION

Covering a large proportion of the burden of non-communicable diseases in adults, we systematically compared the overlap of polygenic risk and different types of family history, showing that they provide independent and complementary information of inherited disease susceptibility in all 24 studied diseases. PRS explained on average 10% of the effect of FH_1st_, but FH_1st_ only 3% of the PRSs, and the PRSs were independent of both early- and late-onset family history. The PRS estimates stratified risk similarly in individuals with and without positive FH: a high PRS conferred a considerably elevated risk, whereas a low PRS compensated for the effect of FH.

To our knowledge, no previous studies have systematically compared how FH and PRS overlap and complement each other across common diseases. Our results are in line with previous disease-specific reports observing at most a modest attenuation in the effect of FH adjusting for PRS in cardiometabolic diseases, cancers, and depression.^11,15,17-27^ We extend these by a systematic comparison across 24 common diseases, using genome-wide PRSs generated with uniform methodology, by measuring FH uniformly through nationwide healthcare registries, and by leveraging genetic relatedness. Our results show that effects of FH and polygenic risk scores are independent, indicating that these measures complement each other for assessment of inherited disease risk. Compared to prevention guidelines that do not recommend use of PRS when FH is available,^3^ these results provide important novel data supporting the use of PRS for improving risk assessment of several diseases with major public health importance.

The largely independent effects have several potential explanations. In addition to capturing shared DNA, FH measures non-genetic exposures and behaviors shared by families. In contrast, PRSs capture each person’s unique combinations of common, disease-associated genetic variants, including genetic risk variation not shared by the relatives. PRSs can be measured in any phase of life, whereas FH relies on disease events having actualized in relatives with utility in late-onset diseases. FH also assigns a similar risk for all relatives of the same degree, despite everyone carrying a unique set of genetic variants measurable through PRSs. Our observation of independent effects is also in line with earlier reports showing the importance of FH of breast and ovarian cancers in *BRCA1* and *BRCA2* mutation carriers.^34^

Genetic information is typically considered in clinical care only when evidence-based prevention strategies to attenuate risk are available.^35^ For instance, risk assessment of cancers has long tradition of comprehensive ascertainment of FH to identify familial clustering^36^ when targeted interventions and screening tools are available.^2,37^ Our results indicate that PRSs could be used to refine risk assessment of breast, prostate, and colorectal cancer, even when information about FH is available. In glaucoma, a high PRS and FH had equal and largely independent effects, but only FH is currently used for assessing risk of glaucoma in patients with ocular hypertension.^38^ The risk of coronary artery disease and type 2 diabetes can be decreased by lifestyle interventions and medications, and FH is commonly used for assessing their risk.^3,39^ For both diseases, we observed larger effects for high PRS than for FH. Moreover, a high PRS may identify individuals more likely to benefit from preventive treatments: for coronary artery disease, a high PRS can result in higher relative efficacy of statins and disclosing PRS risk together with traditional risk factors can motivate lifestyle changes.^40-42^ In contrast, stroke PRSs and FH show lower effect sizes than other cardiovascular diseases, likely owing to the heterogeneity of the disease and differing etiological patterns of stroke subtypes.^43,44^

This study has multiple strengths. FH was assessed systematically and comprehensively by using linkages to high-quality nationwide registries, including hospital discharges, causes-of-death, and medication reimbursement registries, overcoming several limitations of self-reported FH such as recall bias, sensitivity to wording, and inter-individual differences in knowledge about FH. ^5,6,45^ We report effects of FH for disorders challenging to capture precisely from self-reported data, such as alcohol use disorder and atrial fibrillation, and show effects for diseases less studied in the field of PRSs including glaucoma and hypothyroidism. Moreover, we looked at the effect of a dichotomized PRS for comparison purposes and applying PRSs on the continuous scale would provide further risk stratification as unlike FH, extremes of PRSs can also be used to identify individuals at particularly high or low risk. FinnGen’s wide age range is a key strength of the study, allowing systematic comparison of polygenic risk and FH across 24 diseases. Our results are also supported by quantitative genetic theory.^46,47^ Average concordances of a high PRS among first- and second-degree relatives was 33.7% and 19.8%, in line with estimates on cardiometabolic diseases in UK Biobank,^48^ and in agreement theoretically derived concordance estimates of 32.4% and 19.3%.^47^ Moreover, the study provides catalogue of risk estimates for both FH of disease and PRSs in a large-scale biobank study.

The study was limited to individuals of European ancestry among whom current PRSs have the highest utility.^49^ Although our recording of FH_1st_ and FH_2nd_ was primarily based on only one relative, FH_P_ contained both parents, and FH estimates are well in line with earlier reports from epidemiological cohorts and large registry studies (**Supplementary Table 9**). Although the various registries are efficient in capturing cases, milder disease forms such as mild osteoarthritis or atopic dermatitis may remain uncaptured. Similarly, common conditions such as depression or alcohol use disorder are often underreported unless severe or contributing to somatic pathologies. With over half of the study participants in the dataset ascertained from hospital biobanks or disease cohorts, the data is somewhat enriched in cases, resulting in cumulative incidences may not be fully generalizable to the population.

In conclusion, we studied the interplay of family history and genome-wide PRSs, systematically comparing effects across 24 common diseases. The effects of family history and PRS were largely independent, and the pattern was observed across the diseases. We demonstrate that polygenic risk and family history are not interchangeable measures of genetic susceptibility. Instead, they provide complementary information, bringing opportunities for a more comprehensive way of assessing inherited risk. A PRS can be calculated early of life to serve as risk indicator in individuals without family history of disease, while providing effective risk stratification also among individuals with positive family history.

## Acknowledgements

We thank the researchers who have openly shared the GWAS summary statistics that were used in this study to generate PRSs. We would like to thank Mervi Aavikko and Risto Kajanne for management assistance. The FinnGen project is funded by two grants from Business Finland (HUS 4685/31/2016 and UH 4386/31/2016) and the following industry partners: AbbVie Inc., AstraZeneca UK Ltd, Biogen MA Inc., Bristol Myers Squibb, Genentech Inc., Merck Sharp & Dohme Corp, Pfizer Inc., GlaxoSmithKline Intellectual Property Development Ltd., Sanofi US Services Inc., Maze Therapeutics Inc., Janssen Biotech Inc, and Novartis Pharma AG. Following biobanks are acknowledged for delivering biobank samples to FinnGen: Auria Biobank (www.auria.fi/biopankki), THL Biobank (www.thl.fi/biobank), Helsinki Biobank (www.helsinginbiopankki.fi), Biobank Borealis of Northern Finland (https://www.ppshp.fi/Tutkimus-ja-opetus/Biopankki/Pages/Biobank-Borealis-briefly-in-English.aspx), Finnish Clinical Biobank Tampere (www.tays.fi/en-US/Research_and_development/Finnish_Clinical_Biobank_Tampere), Biobank of Eastern Finland (www.ita-suomenbiopankki.fi/en), Central Finland Biobank (www.ksshp.fi/fi-FI/Potilaalle/Biopankki), Finnish Red Cross Blood Service Biobank (www.veripalvelu.fi/verenluovutus/biopankkitoiminta) and Terveystalo Biobank (www.terveystalo.com/fi/Yritystietoa/Terveystalo-Biopankki/Biopankki/). All Finnish Biobanks are members of BBMRI.fi infrastructure (www.bbmri.fi) and FINBB biobank cooperative (https://finbb.fi/) is the coordinator of the BBMRI-ERIC operations in Finland covering all Finnish biobanks.

This work has been supported by Academy of Finland (grant number 331671 to N.M., 285380 to S.R., 128650 to A.P., 308248 to J.K.); Academy of Finland Center of Excellence in Complex Disease Genetics (grant number 312062 to S.R., 312074 to A.P., 312073 to J.K.); European Union’s Horizon 2020 research and innovation programme under grant agreement No 101016775; the Sigrid Jusélius Foundation (to S.R. and A.P.); University of Helsinki HiLIFE Fellow grants 2017-2020 (to S.R.); The Finnish Innovation Fund Tekes (grant number 2273/31/2017 to E.W.); Foundation and the Horizon 2020 Research and Innovation Programme (grant number 667301 (COSYN) to A.P.); JVL was supported by Academy of Finland (grant number 311492) and Helsinki Institute of Life Science (H970).

## Data availability

The FinnGen data may be accessed through Finnish Biobanks’ FinBB portal (web link: www.finbb.fi). Download links for the GWAS summary statistics used for constructing are provided in Supplementary Table 1. The weights for our polygenic risk scores will be made are available at PGS Catalog, with the submitted data embargoed until publication and PGS Catalog identifiers reported in the final version of the manuscript.

## Code availability

The full genotyping and imputation protocol for FinnGen is described at https://doi.org/10.17504/protocols.io.xbgfijw. The PRS-CS pipeline in FinnGen is described at https://github.com/FINNGEN/CS-PRS-pipeline.

## Author contributions

N.M., J.V.L, and S.R. conceived and designed the study. N.M. and P.D.B.P carried out the statistical and computational analyses with advice from S.R. and J.V.L. Quality control of the data was carried out by N.M. and P.D.B.P. All authors provided critical inputs to interpretation of the data. The manuscript was written and revised by N.M. and S.R., with comments from all of the co-authors. All co-authors have approved the final version of the manuscript.

