## Supplementary Methods and Figures for "Systematic comparison of family history and polygenic risk across 24 common diseases"

Identification of cases included harmonization of diagnoses according to different revisions of International Statistical Classification of Diseases (ICD-8/9/10) (**Supplementary Table 12**). Registries used include the hospital discharge registry (available from 1968-), the Finnish Cancer Registry (1953-), causes of death registry (1964-), and the medication reimbursement and medication purchases registries (1964- and 1995-), both administered by the Social Insurance Institute of Finland. Age at disease onset was defined as the age at the first healthcare contact with the disease. Age at death from the causes of death registry as recorded on the death certificate and checked by Statistics Finland. Parental cause of death ($FH_P$) was defined as at least one parent having the disease as a cause of death. Prostate cancer was studied only in men and breast cancer only in women.

#### Genotyping and imputation

FinnGen samples were genotyped with Illumina and Affymetrix arrays (Illumina Inc., San Diego, and Thermo Fisher Scientific, Santa Clara, CA, USA), and genotype calls were made with the GenCall or zCall (for Illumina) and the AxiomGT1 algorithm for Affymetrix data. Individuals with ambiguous gender, high genotype missingness (>5%), excess heterozygosity (+-4SD) and non-Finnish ancestry were excluded, as well as all variants with high missingness (>2%), low Hardy–Weinberg equilibrium p-value (<1e-6) and minor allele count (MAC < 3). Array data pre-phasing was carried out with Eagle 2.3.5[1] with the number of conditioning haplotypes set to 20,000. Genotype imputation was done using the population-specific SISu v3 imputation reference with 3,775 high-coverage (25-30x) whole-genome sequences in Finns, described in detail at https://doi.org/10.17504/protocols.io.xbgfijw.

#### Polygenic risk scores

We used the PRS-CS-auto algorithm, which learns the model's global scaling parameter $\phi$ from the data, performing well with large datasets.[2] The PRSs for autosomes were calculated using PLINK v2.00a2.3LM, by calculating the weighted sum of risk alleles for each variant with the parameter *--score* applied on a genotype file (all chromosomes combined) which had been filtered to 1,194,526 HapMap3 variants, as recommended for PRS-CS. The mean number of variants included in the PRSs was 1,059,217. Details on the GWASs used as the input for the PRSs are available in **Supplementary Table 4**. We observed very little correlation between the PRSs (**Supplementary Figure 9**). To avoid overfitting of effects, the individuals potentially overlapping with the GWASs were excluded from all analyses based on genotyping array and cohort information (**Supplementary Tables 4 and 11**). As we are unable to identify the exact individuals overlapping, we chose to use this conservative exclusion approach.

#### Inferring relatedness

To define first-degree family history ($FH_{1st}$), we inferred first-degree relatedness from genotypes based on 173,907 independent linkage disequilibrium (LD)-pruned common variants 9 (PLINK parameters --snps-only --chr 1-22 --max-alleles 2 --maf 0.01 --indep-pairwise 500.0 50.0 0.15). The LD pruning was done using variants with INFO >0.9. Using KING v2.2.4[3], pairs of first-degree relatives were identified with a kinship coefficient between 0.177 and 0.354, and pairs of second-degree relatives with a kinship coefficient between 0.0884 and 0.177. To avoid individuals appearing multiple times on either side of the regression equation which would violate the assumption of independence of observations, we performed several steps of random exclusions and exclusions of cohorts predominantly family-based ascertainment (**Supplementary Figure 1**). We inferred the risk for the individual born later and the individual born earlier was chosen as the relative (random choice for dizygotic twins). The PRS effect sizes were similar in the full data and in those with a first-degree relative in the dataset (**Supplementary Figure 10**).

#### Ethics statement

Patients and control subjects in FinnGen provided informed consent for biobank research, based on the Finnish Biobank Act. Alternatively, separate research cohorts, collected prior the Finnish Biobank Act came into effect (in September 2013) and start of FinnGen (August 2017), were collected based on study-specific consents and later transferred to the Finnish biobanks after approval by Fimea (Finnish Medicines Agency), the National Supervisory Authority for Welfare and Health. Recruitment protocols followed the biobank protocols approved by Fimea. The Coordinating Ethics Committee of the Hospital District of Helsinki and Uusimaa (HUS) statement number for the FinnGen study is Nr HUS/990/2017.

The FinnGen study is approved by Finnish Institute for Health and Welfare (permit numbers: THL/2031/6.02.00/2017, THL/1101/5.05.00/2017, THL/341/6.02.00/2018, THL/2222/6.02.00/ 2018, THL/283/6.02.00/2019, THL/1721/5.05.00/2019, THL/1524/5.05.00/2020, and THL/2364/ 14.02/2020), Digital and population data service agency (permit numbers: VRK43431/2017-3, VRK/6909/2018-3, VRK/4415/2019-3), the Social Insurance Institution (permit numbers: KELA 58/522/2017, KELA 131/522/2018, KELA 70/522/2019, KELA 98/522/2019, KELA 138/522/2019, KELA 2/522/2020, KELA 16/522/2020, Findata THL/2364/14.02/2020 and Statistics Finland (permit numbers: TK-53-1041-17 and TK/143/07.03.00/2020 (earlier TK-53-90-20).

The Biobank Access Decisions for FinnGen samples and data utilized in FinnGen Data Freeze 7 include: THL Biobank BB2017_55, BB2017_111, BB2018_19, BB_2018_34, BB_2018_67, BB2018_71, BB2019_7, BB2019_8, BB2019_26, BB2020_1, Finnish Red Cross Blood Service Biobank 7.12.2017, Helsinki Biobank HUS/359/2017, Auria Biobank AB17-5154 and amendment #1 (August 17 2020), Biobank Borealis of Northern Finland_2017_1013, Biobank of Eastern Finland 1186/2018 and amendment 22 § /2020, Finnish Clinical Biobank Tampere MH0004 and amendments (21.02.2020 & 06.10.2020), Central Finland Biobank 1-2017, and Terveystalo Biobank STB 2018001.

**Supplementary Figure 1.** Study flowchart describing generation of datasets used for the definitions first-degree family history and parental cause of death. Having inferred kinship, the initial data structure allows individuals to appear multiple times in the dataset within different pairs. The main processing steps after this involved random exclusions, to individuals appearing multiple times on either side of the regression equation, which would violate the assumption of independence of observations. Similar steps were performed for second-degree family history as for first degree family history, starting from 118,992 pairs of individuals with a second-degree relative in the dataset and resulting in 47,154 individuals for analysis of second-degree family history as a risk factor. For breast cancer, we studied only pairs of women (15,281 individuals, parent-offspring relationship in 7,770; full-sibling relationship in 7,511). For prostate cancer, we studied only pairs of men (9,473 individuals, parent-offspring relationship in 3,932; full-sibling relationship in 5,541).

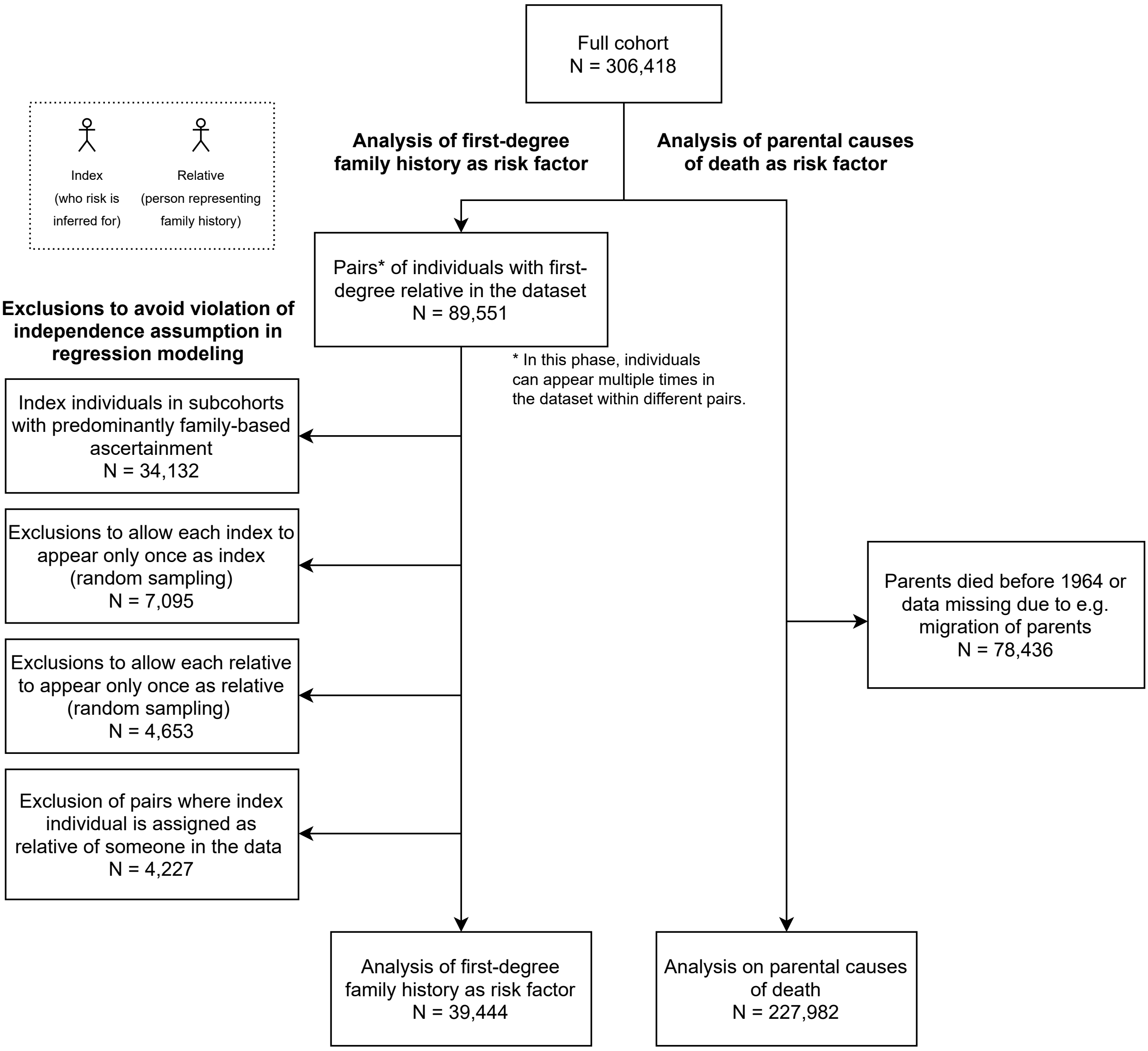

**Supplementary Figure 2.** Prevalence of first-degree family history ($FH_{1st}$) by deciles of polygenic risk score (PRS).

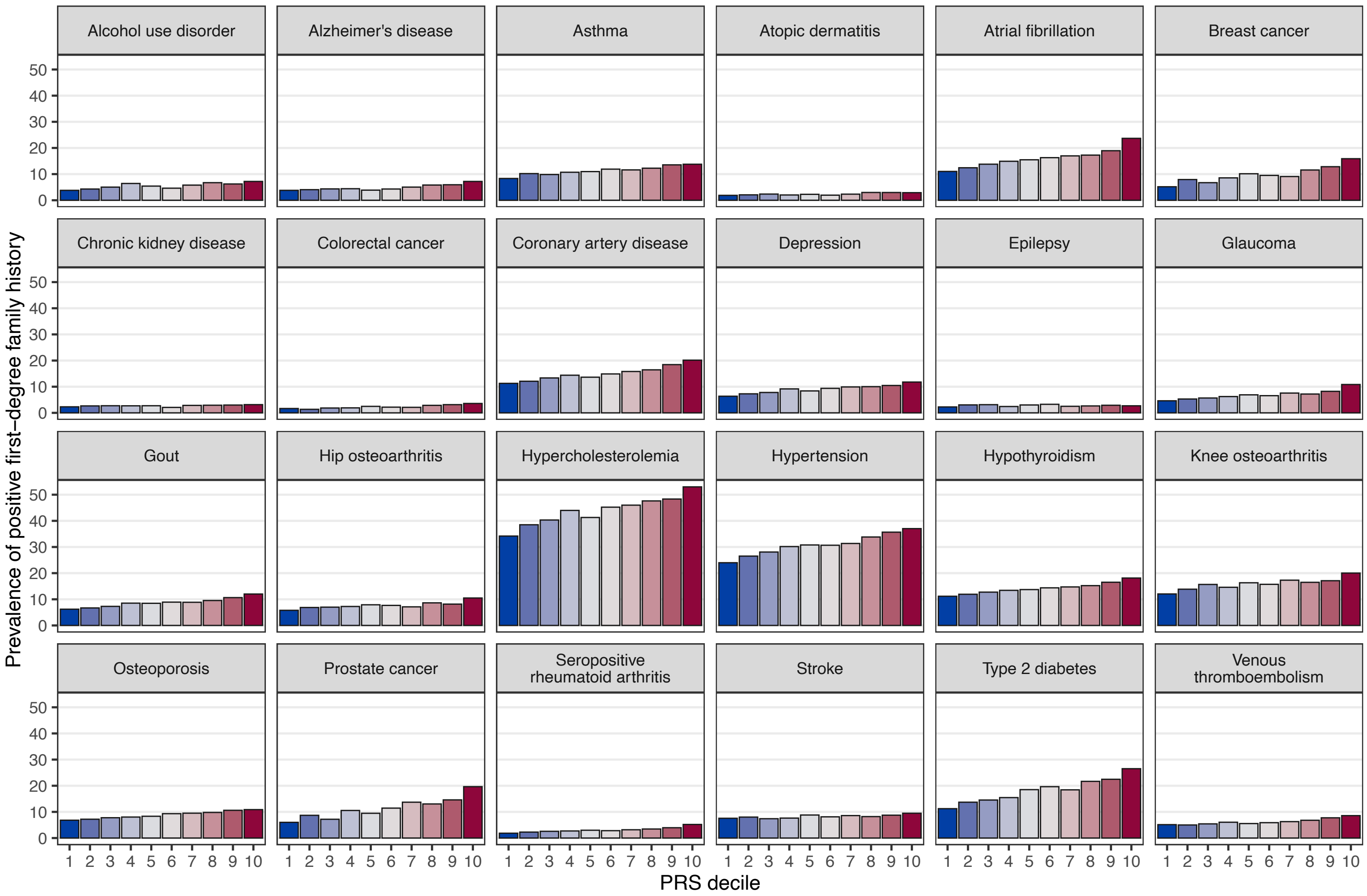


Total N = 39,444, N = 15,281 for breast cancer, N = 9,473 for prostate cancer.

**Supplementary Figure 3.** Cross-adjustment effects for family history and polygenic risk score (PRS) with PRS categorized. The impact of adjusting the PRS effect with first-degree family history **(**FH$_{1st}$, **panel A)** and vice versa **(panel B)**. The diamonds represent the unadjusted effects and the squares the adjusted effects. The PRS effect size compares individuals in the top decile of the PRS distribution to the rest.

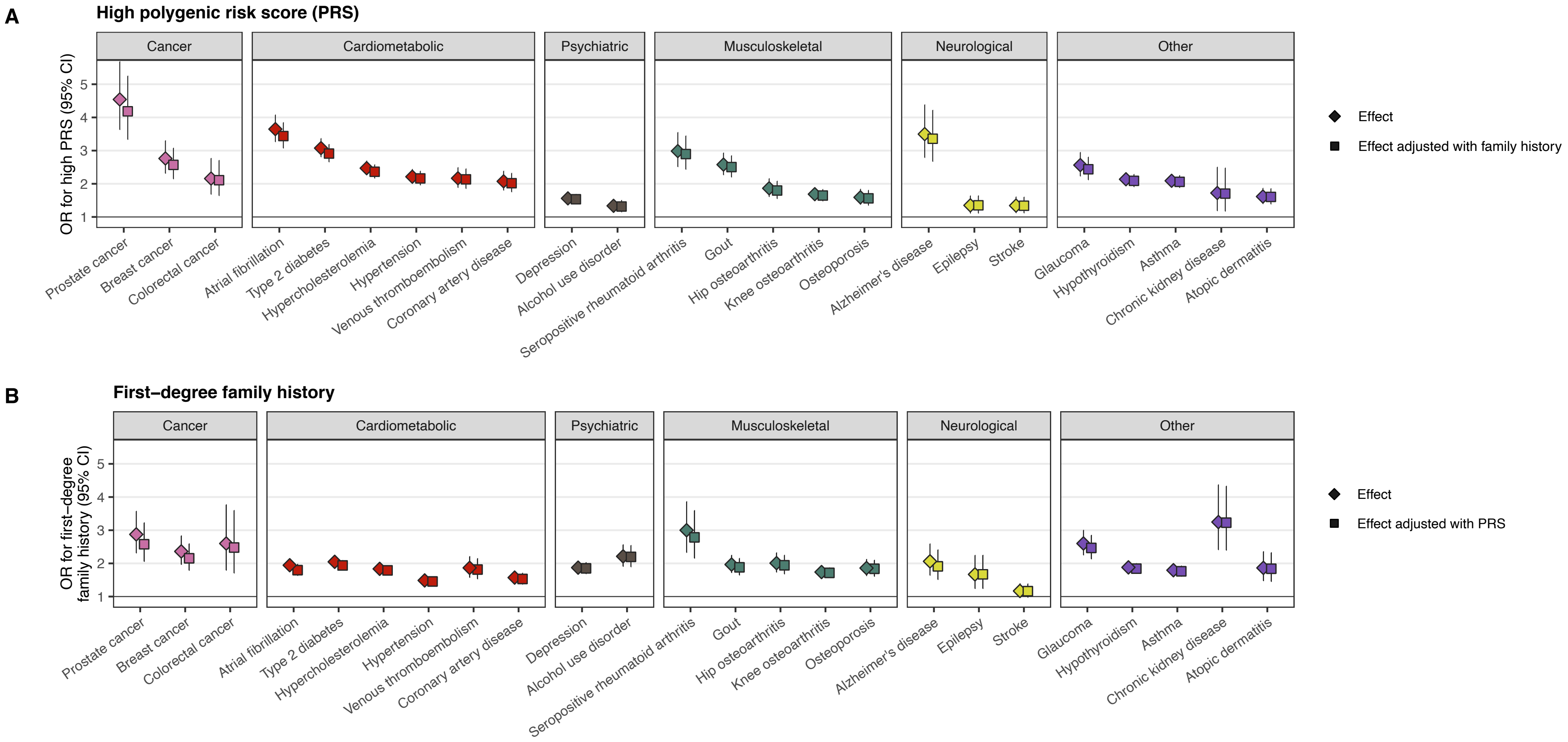


Total N = 39,444, N = 15,281 for breast cancer, N = 9,473 for prostate cancer. Odds ratios (OR) were obtained from logistic regression models adjusted for sex (except for breast and prostate cancer), birth year, genotyping array, cohort, and the first ten genetic principal components of ancestry.

**Supplementary Figure 4.** Interaction analysis between first-degree family history ($FH_{1st}$, **panel A**) or parental causes of death ($FH_P$, **panel B**) and respective polygenic risk scores (PRS), displaying the p-value for the interaction term on the y-axis. **Panel C** shows the results of **panel B** adjusting also for birth year of the relative. The PRSs were scaled to zero mean and unit variance and handled as continuous variables in the interaction analysis. Statistical significance set at a p-value threshold of 0.0013 (Bonferroni-correction for 24+15 tests) represented by the red line. We did not observe systematic evidence of interactions.

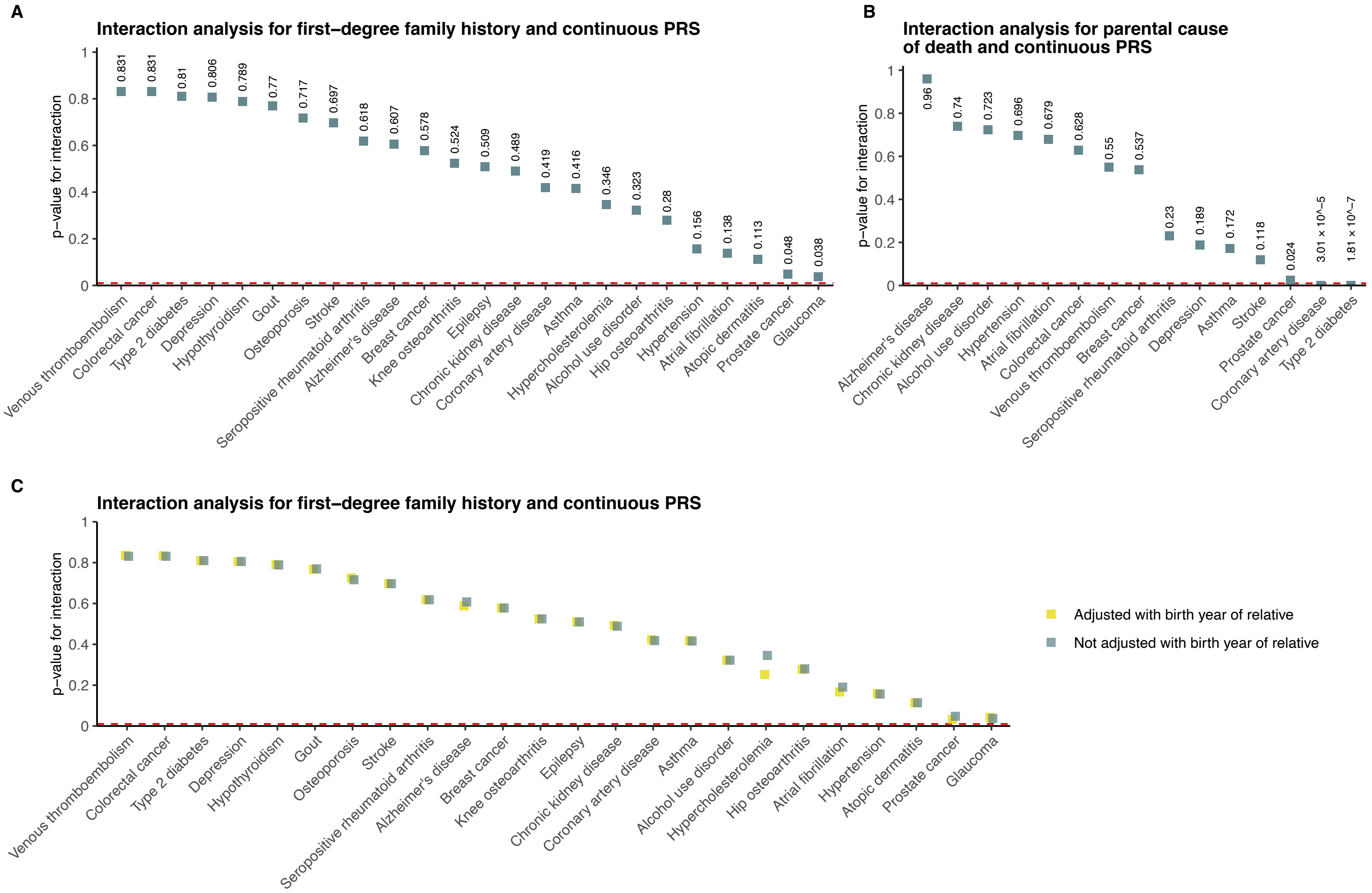


Total N = 39,444, N = 15,281 for breast cancer, N = 9,473 for prostate cancer. The logistic regression models were adjusted for sex (except for breast and prostate cancer), birth year, genotyping array, cohort, and the first ten genetic principal components of ancestry.

**Supplementary Figure 5.** The impact of polygenic risk on disease risk in individuals with positive family history across a broad set of diseases. Analyses were performed for diseases with an OR >2 for high PRS in Figure 2 (15/24 diseases), and over 10 cases in each subgroup, excluding Alzheimer's disease due to its average onset late in life. The survival curves show cumulative incidences for individuals with positive first-degree family ($FH_{1st}$), stratified by level of polygenic risk score (PRS). High PRS was defined as top decile of the PRS distribution and low PRS as the bottom tertile of the PRS distribution.

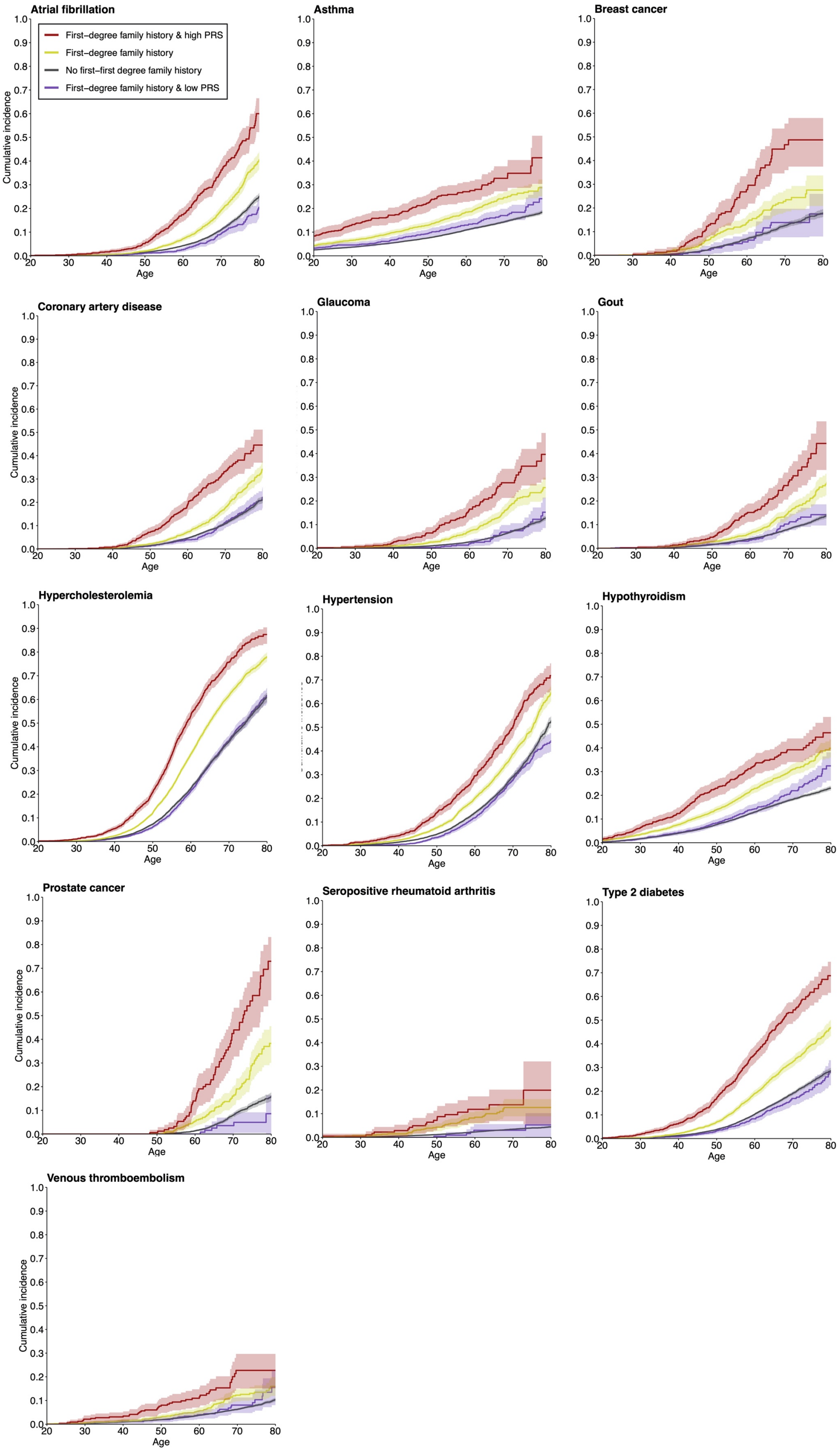


Total N = 39,444, N = 15,281 for breast cancer, N = 9,473 for prostate cancer.

**Supplementary Figure 6.** Impact of the level of polygenic risk score (PRS) on cumulative incidence of disease in individuals with negative (dashed lines) and positive (solid line) first-degree family ($FH_{1st}$). High PRS was defined as top decile of the PRS distribution and low PRS as the bottom tertile of the PRS distribution. A low PRS compensated for the impact of positive $FH_{1st}$, whereas individuals with a combination of high PRS and positive $FH_{1st}$ had a particularly high risk. The figure shows results for the five diseases with the largest effect sizes for PRS, and for breast and prostate cancer.

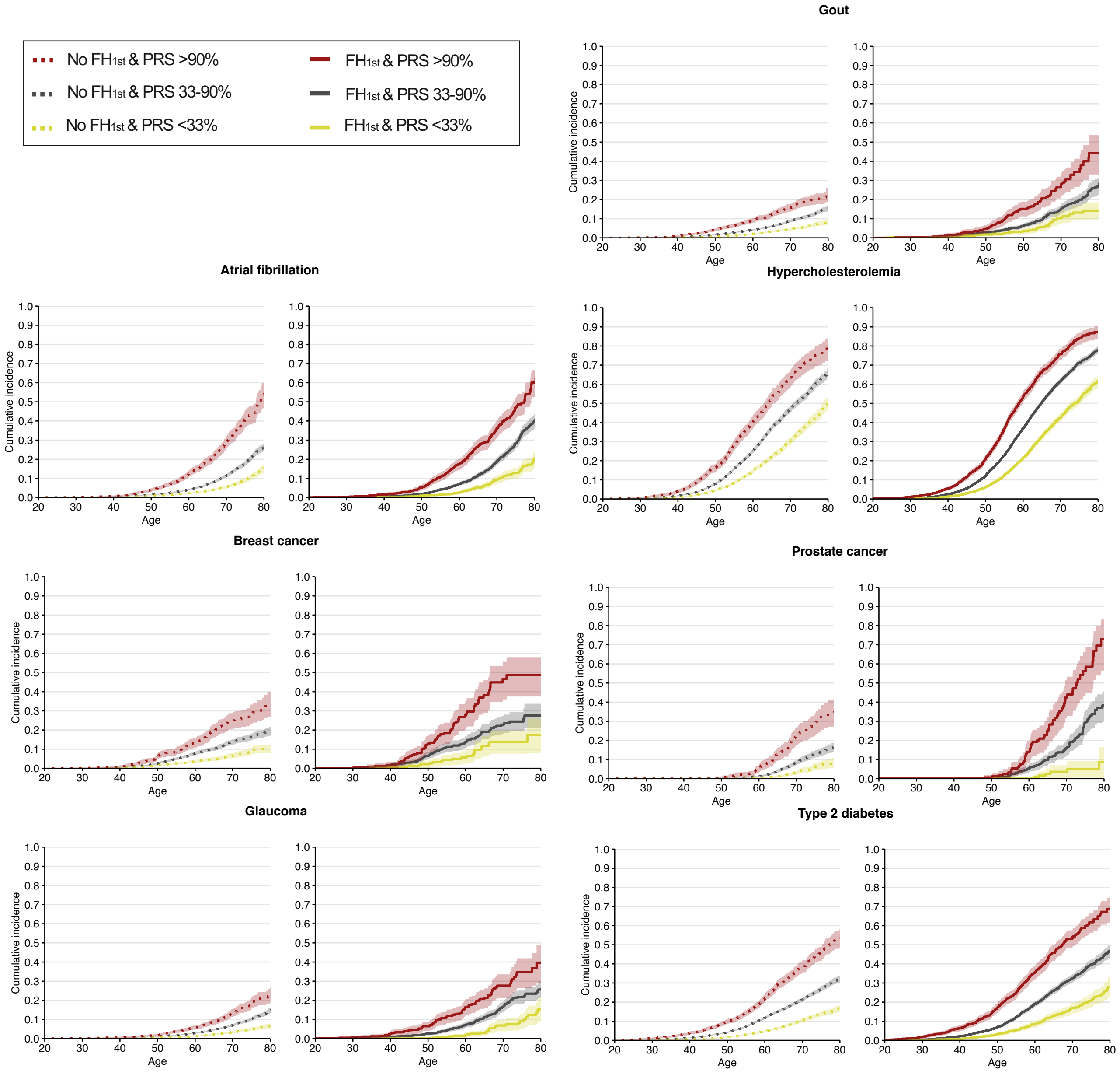


Total N = 39,444, N = 15,281 for breast cancer, N = 9,473 for prostate cancer.

**Supplementary Figure 7.** Concordance of a high polygenic risk score (PRS; defined as top 10% of the distribution) among first-degree relatives (**panel A**) and among second-degree relatives (**panel B**). The horizontal lines denote the theoretically derived concordance estimates of 32.4% (**panel A**) and 19.3% (**panel B**) calculated based on reference 47 for first-degree relatives using a high PRS defined as the top 10% of the distribution.

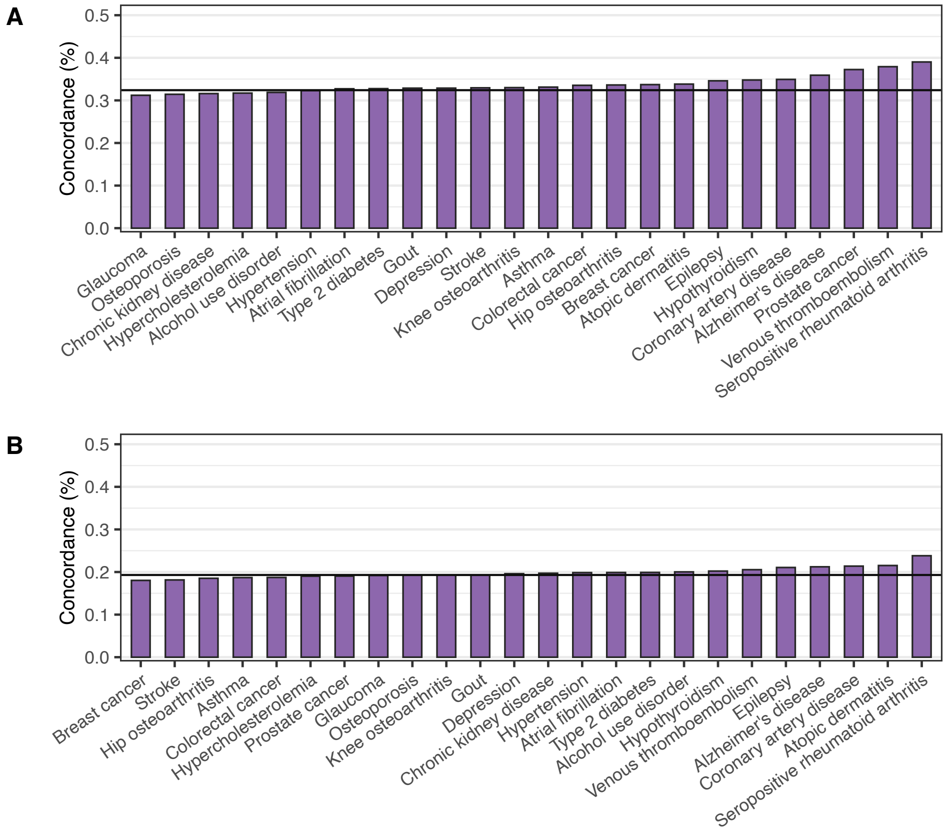


Sample sizes: panel A total N = 39,444, N = 15,281 for breast cancer, N = 9,473 for prostate cancer; panel B second -degree family history total N = 47,154, N = 18,973 for breast cancer, N = 12,355 for prostate cancer.

**Supplementary Figure 8.** Concordance of a high polygenic risk score (PRS; defined as top 10% of the distribution) among first-degree relatives (**panel A**) and among second-degree relatives (**panel B**) stratifying by the respective family history status.

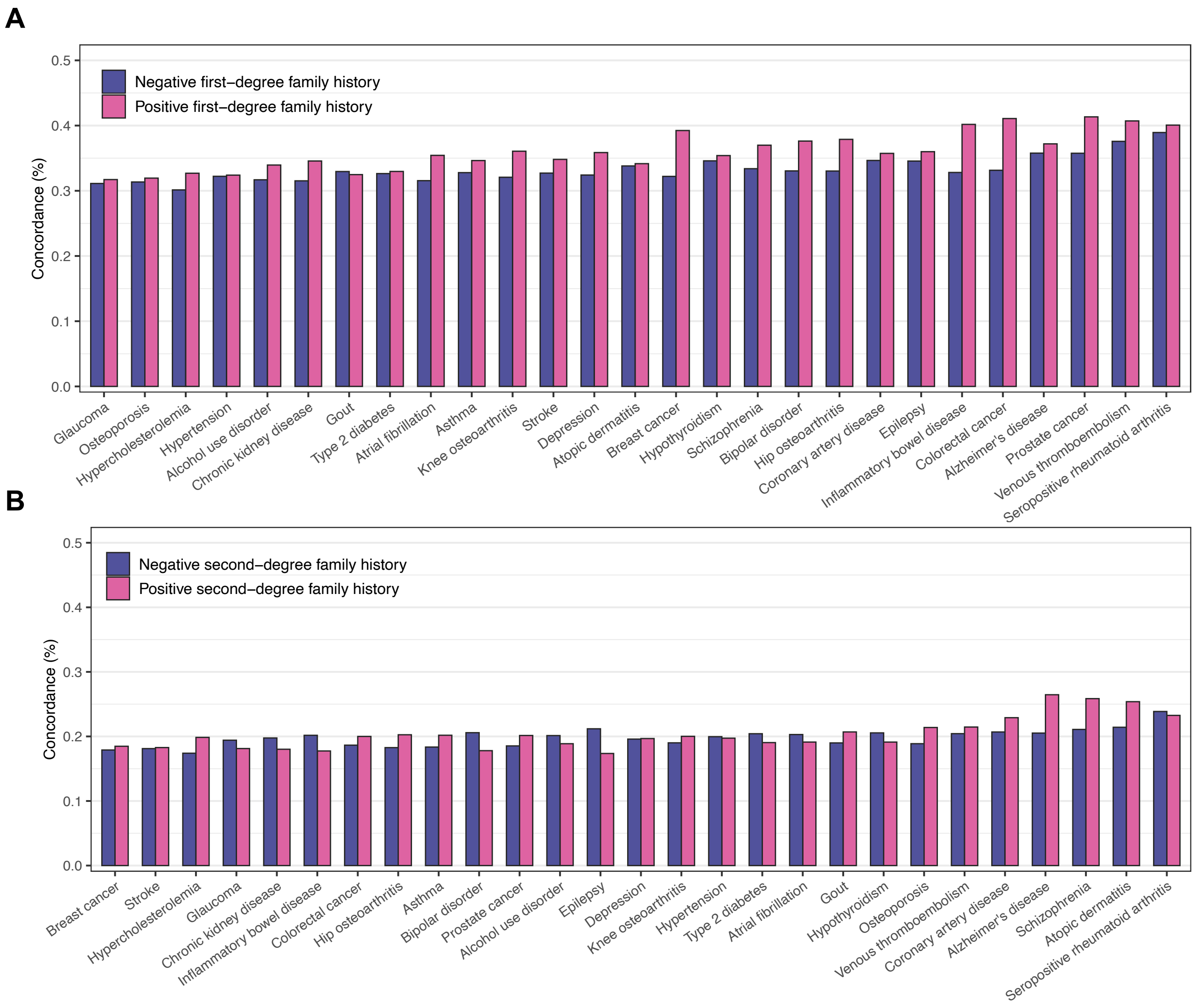


Sample sizes: panel A total N = 39,444, N = 15,281 for breast cancer, N = 9,473 for prostate cancer; panel B second -degree family history total N = 47,154, N = 18,973 for breast cancer, N = 12,355 for prostate cancer.

**Supplementary Figure 9.** Pearson correlation of the 24 disease-specific polygenic risk scores (PRS), assessed on the continuous scale.

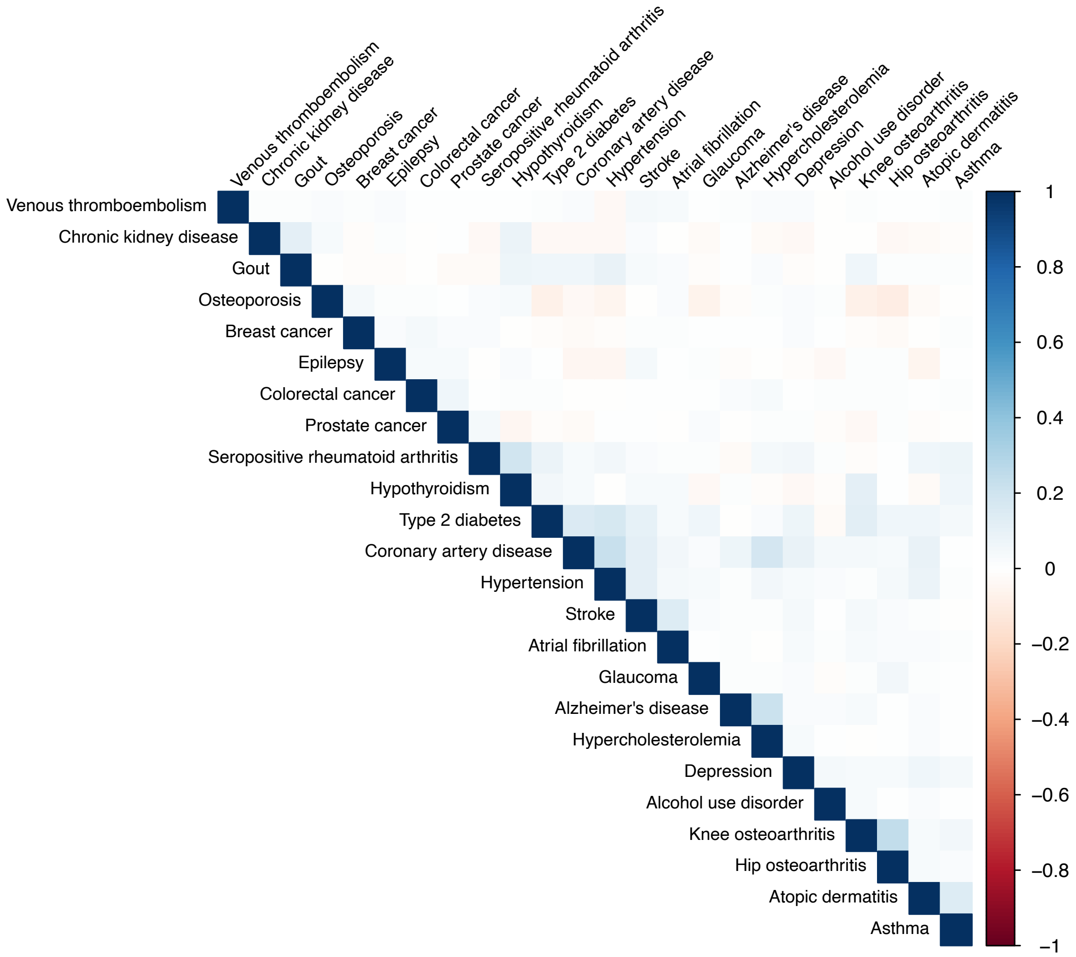

**Supplementary Figure 10.** Effect sizes for a high polygenic risk score (PRS; defined as top 10% of the distribution), comparing individuals in the full data (N = 306,418) and the dataset used for analyses on first-degree relatives (N = 39,444). The PRS effect sizes were similar in both.

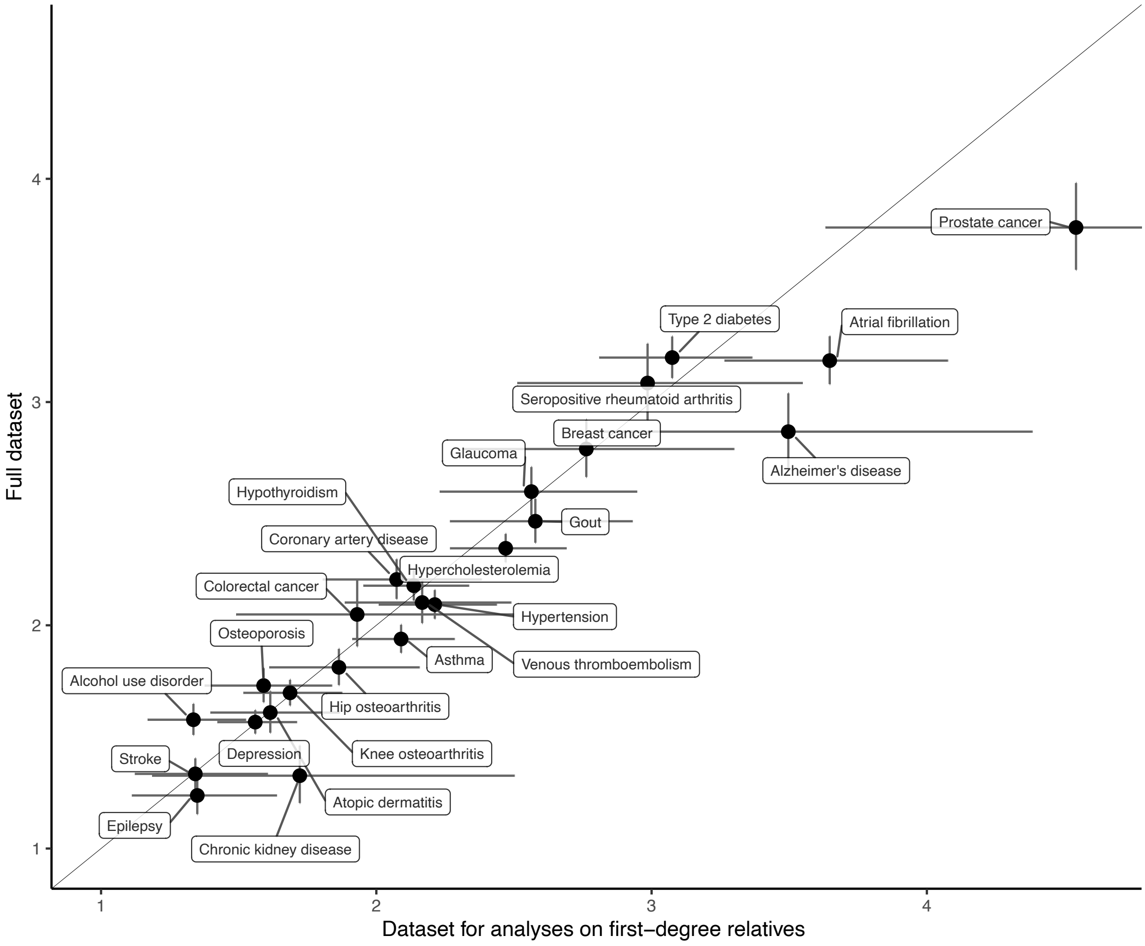

### FinnGen


| Full Name | Affiliation |
|---|---|
| Aarno Palotie | Institute for Molecular Medicine Finland (FIMM), HiLIFE, University of Helsinki, Helsinki, Finland; Broad Institute of MIT and Harvard; Massachusetts General Hospital |
| Mark Daly | Institute for Molecular Medicine Finland (FIMM), HiLIFE, University of Helsinki, Helsinki, Finland; Broad Institute of MIT and Harvard; Massachusetts General Hospital |
| Bridget Riley-Gills | Abbvie, Chicago, IL, United States |
| Howard Jacob | Abbvie, Chicago, IL, United States |
| Dirk Paul | Astra Zeneca, Cambridge, United Kingdom |
| Athena Matakidou | Astra Zeneca, Cambridge, United Kingdom |
| Adam Platt | Astra Zeneca, Cambridge, United Kingdom |
| Heiko Runz | Biogen, Cambridge, MA, United States |
| Sally John | Biogen, Cambridge, MA, United States |
| George Okafo | Boehringer Ingelheim, Ingelheim am Rhein, Germany |
| Nathan Lawless | Boehringer Ingelheim, Ingelheim am Rhein, Germany |
| Robert Plenge | Bristol Myers Squibb, New York, NY, United States |
| Joseph Maranville | Bristol Myers Squibb, New York, NY, United States |
| Mark McCarthy | Genentech, San Francisco, CA, United States |
| Julie Hunkapiller | Genentech, San Francisco, CA, United States |
| Margaret G. Ehm | GlaxoSmithKline, Collegeville, PA, United States |
| Kirsi Auro | GlaxoSmithKline, Espoo, Finland |
| Simonne Longerich | Merck, Kenilworth, NJ, United States |
| Caroline Fox | Merck, Kenilworth, NJ, United States |
| Anders Mälarstig | Pfizer, New York, NY, United States |
| Katherine Klinger | Translational Sciences, Sanofi R&D, Framingham, MA, USA |
| Deepak Raipal | Translational Sciences, Sanofi R&D, Framingham, MA, USA |
| Eric Green | Maze Therapeutics, San Francisco, CA, United States |
| Robert Graham | Maze Therapeutics, San Francisco, CA, United States |
| Robert Yang | Janssen Biotech, Beerse, Belgium |
| Chris O´Donnell | Novartis Institutes for BioMedical Research, Cambridge, MA, United States |
| Tomi P. Mäkelä | HiLIFE, University of Helsinki, Finland, Finland |
| Jaakko Kaprio | Institute for Molecular Medicine Finland (FIMM), HiLIFE, University of Helsinki, Helsinki, Finland |
| Petri Virolainen | Auria Biobank / University of Turku / Hospital District of Southwest Finland, Turku, Finland |
| Antti Hakanen | Auria Biobank / University of Turku / Hospital District of Southwest Finland, Turku, Finland |
| Terhi Kilpi | THL Biobank / Finnish Institute for Health and Welfare (THL), Helsinki, Finland |
| Markus Perola | THL Biobank / Finnish Institute for Health and Welfare (THL), Helsinki, Finland |
| Jukka Partanen | Finnish Red Cross Blood Service / Finnish Hematology Registry and Clinical Biobank, Helsinki, Finland |
| Anne Pitkäranta | Helsinki Biobank / Helsinki University and Hospital District of Helsinki and Uusimaa, Helsinki |
| Juhani Junttila | Northern Finland Biobank Borealis / University of Oulu / Northern Ostrobothnia Hospital District, Oulu, Finland |
| Raisa Serpi | Northern Finland Biobank Borealis / University of Oulu / Northern Ostrobothnia Hospital District, Oulu, Finland |
| Tarja Laitinen | Finnish Clinical Biobank Tampere / University of Tampere / Pirkanmaa Hospital District, Tampere, Finland |
| Veli-Matti Kosma | Biobank of Eastern Finland / University of Eastern Finland / Northern Savo Hospital District, Kuopio, Finland |
| Jari Laukkanen | Central Finland Biobank / University of Jyväskylä / Central Finland Health Care District, Jyväskylä, Finland |
| Marco Hautalahti | FINBB - Finnish biobank cooperative |
| Outi Tuovila | Business Finland, Helsinki, Finland |
| Raimo Pakkanen | Business Finland, Helsinki, Finland |
| Jeffrey Waring | Abbvie, Chicago, IL, United States |
| Bridget Riley-Gillis | Abbvie, Chicago, IL, United States |
| Fedik Rahimov | Abbvie, Chicago, IL, United States |
| Ioanna Tachmazidou | Astra Zeneca, Cambridge, United Kingdom |
| Chia-Yen Chen | Biogen, Cambridge, MA, United States |
| Heiko Runz | Biogen, Cambridge, MA, United States |
| Zhihao Ding | Boehringer Ingelheim, Ingelheim am Rhein, Germany |
| Marc Jung | Boehringer Ingelheim, Ingelheim am Rhein, Germany |
| Shameek Biswas | Bristol Myers Squibb, New York, NY, United States |
| Rion Pendergrass | Genentech, San Francisco, CA, United States |
| Julie Hunkapiller | Genentech, San Francisco, CA, United States |
| Margaret G. Ehm | GlaxoSmithKline, Collegeville, PA, United States |
| David Pulford | GlaxoSmithKline, Stevenage, United Kingdom |
| Neha Raghavan | Merck, Kenilworth, NJ, United States |
| Adriana Huertas-Vazquez | Merck, Kenilworth, NJ, United States |
| Jae-Hoon Sul | Merck, Kenilworth, NJ, United States |
| Anders Mälarstig | Pfizer, New York, NY, United States |
| Xinli Hu | Pfizer, New York, NY, United States |
| Katherine Klinger | Translational Sciences, Sanofi R&D, Framingham, MA, USA |

| | |
|---|---|
| Robert Graham | Maze Therapeutics, San Francisco, CA, United States |
| Eric Green | Maze Therapeutics, San Francisco, CA, United States |
| Sahar Mozaffari | Maze Therapeutics, San Francisco, CA, United States |
| Dawn Waterworth | Janssen Research & Development, LLC, Spring House, PA, United States |
| Nicole Renaud | Novartis Institutes for BioMedical Research, Cambridge, MA, United States |
| Ma´en Obeidat | Novartis Institutes for BioMedical Research, Cambridge, MA, United States |
| Samuli Ripatti | Institute for Molecular Medicine Finland (FIMM), HiLIFE, University of Helsinki, Helsinki, Finland |
| Johanna Schleutker | Auria Biobank / Univ. of Turku / Hospital District of Southwest Finland, Turku, Finland |
| Markus Perola | THL Biobank / Finnish Institute for Health and Welfare (THL), Helsinki, Finland |
| Mikko Arvas | Finnish Red Cross Blood Service / Finnish Hematology Registry and Clinical Biobank, Helsinki, Finland |
| Olli Carpén | Helsinki Biobank / Helsinki University and Hospital District of Helsinki and Uusimaa, Helsinki |
| Reetta Hinttala | Northern Finland Biobank Borealis / University of Oulu / Northern Ostrobothnia Hospital District, Oulu, Finland |
| Johannes Kettunen | Northern Finland Biobank Borealis / University of Oulu / Northern Ostrobothnia Hospital District, Oulu, Finland |
| Arto Mannermaa | Biobank of Eastern Finland / University of Eastern Finland / Northern Savo Hospital District, Kuopio, Finland |
| Katriina Aalto-Setälä | Faculty of Medicine and Health Technology, Tampere University, Tampere, Finland |
| Mika Kähönen | Finnish Clinical Biobank Tampere / University of Tampere / Pirkanmaa Hospital District, Tampere, Finland |
| Jari Laukkanen | Central Finland Biobank / University of Jyväskylä / Central Finland Health Care District, Jyväskylä, Finland |
| Johanna Mäkelä | FINBB - Finnish biobank cooperative |
| Reetta Kälviäinen | Northern Savo Hospital District, Kuopio, Finland |
| Valtteri Julkunen | Northern Savo Hospital District, Kuopio, Finland |
| Hilkka Soininen | Northern Savo Hospital District, Kuopio, Finland |
| Anne Remes | Northern Ostrobothnia Hospital District, Oulu, Finland |
| Mikko Hiltunen | University of Eastern Finland, Kuopio, Finland |
| Jukka Peltola | Pirkanmaa Hospital District, Tampere, Finland |
| Minna Raivio | Hospital District of Helsinki and Uusimaa, Helsinki, Finland |
| Pentti Tienari | Hospital District of Helsinki and Uusimaa, Helsinki, Finland |
| Juha Rinne | Hospital District of Southwest Finland, Turku, Finland |
| Roosa Kallionpää | Hospital District of Southwest Finland, Turku, Finland |
| Juulia Partanen | Institute for Molecular Medicine Finland, HiLIFE, University of Helsinki, Finland |
| Ali Abbasi | Abbvie, Chicago, IL, United States |
| Adam Ziemann | Abbvie, Chicago, IL, United States |
| Nizar Smaoui | Abbvie, Chicago, IL, United States |
| Anne Lehtonen | Abbvie, Chicago, IL, United States |
| Susan Eaton | Biogen, Cambridge, MA, United States |
| Heiko Runz | Biogen, Cambridge, MA, United States |
| Sanni Lahdenperä | Biogen, Cambridge, MA, United States |
| Shameek Biswas | Bristol Myers Squibb, New York, NY, United States |
| Julie Hunkapiller | Genentech, San Francisco, CA, United States |
| Natalie Bowers | Genentech, San Francisco, CA, United States |
| Edmond Teng | Genentech, San Francisco, CA, United States |
| Rion Pendergrass | Genentech, San Francisco, CA, United States |
| Fanli Xu | GlaxoSmithKline, Brentford, United Kingdom |
| David Pulford | GlaxoSmithKline, Stevenage, United Kingdom |
| Kirsi Auro | GlaxoSmithKline, Espoo, Finland |
| Laura Addis | GlaxoSmithKline, Brentford, United Kingdom |
| John Eicher | GlaxoSmithKline, Brentford, United Kingdom |
| Qingqin S Li | Janssen Research & Development, LLC, Titusville, NJ 08560, United States |
| Karen He | Janssen Research & Development, LLC, Spring House, PA, United States |
| Ekaterina Khramtsova | Janssen Research & Development, LLC, Spring House, PA, United States |
| Neha Raghavan | Merck, Kenilworth, NJ, United States |
| Martti Färkkilä | Hospital District of Helsinki and Uusimaa, Helsinki, Finland |
| Jukka Koskela | Hospital District of Helsinki and Uusimaa, Helsinki, Finland |
| Sampsa Pikkarainen | Hospital District of Helsinki and Uusimaa, Helsinki, Finland |
| Airi Jussila | Pirkanmaa Hospital District, Tampere, Finland |
| Katri Kaukinen | Pirkanmaa Hospital District, Tampere, Finland |
| Timo Blomster | Northern Ostrobothnia Hospital District, Oulu, Finland |
| Mikko Kiviniemi | Northern Savo Hospital District, Kuopio, Finland |
| Markku Voutilainen | Hospital District of Southwest Finland, Turku, Finland |
| Mark Daly | Institute for Molecular Medicine, Finland (FIMM), HiLIFE, University of Helsinki, Helsinki, Finland; Broad Institute of MIT and Harvard; Massachusetts General Hospital |
| Ali Abbasi | Abbvie, Chicago, IL, United States |
| Jeffrey Waring | Abbvie, Chicago, IL, United States |
| Nizar Smaoui | Abbvie, Chicago, IL, United States |
| Fedik Rahimov | Abbvie, Chicago, IL, United States |
| Anne Lehtonen | Abbvie, Chicago, IL, United States |
| Tim Lu | Genentech, San Francisco, CA, United States |
| Natalie Bowers | Genentech, San Francisco, CA, United States |

| | |
|---|---|
| Rion Pendergrass | Genentech, San Francisco, CA, United States |
| Linda McCarthy | GlaxoSmithKline, Brentford, United Kingdom |
| Amy Hart | Janssen Research & Development, LLC, Spring House, PA, United States |
| Meijian Guan | Janssen Research & Development, LLC, Spring House, PA, United States |
| Jason Miller | Merck, Kenilworth, NJ, United States |
| Kirsi Kalpala | Pfizer, New York, NY, United States |
| Melissa Miller | Pfizer, New York, NY, United States |
| Xinli Hu | Pfizer, New York, NY, United States |
| Kari Eklund | Hospital District of Helsinki and Uusimaa, Helsinki, Finland |
| Antti Palomäki | Hospital District of Southwest Finland, Turku, Finland |
| Pia Isomäki | Pirkanmaa Hospital District, Tampere, Finland |
| Laura Pirilä | Hospital District of Southwest Finland, Turku, Finland |
| Oili Kaipiainen-Seppänen | Northern Savo Hospital District, Kuopio, Finland |
| Johanna Huhtakangas | Northern Ostrobothnia Hospital District, Oulu, Finland |
| Nina Mars | Institute for Molecular Medicine Finland (FIMM), HiLIFE, University of Helsinki, Helsinki, Finland |
| Ali Abbasi | Abbvie, Chicago, IL, United States |
| Jeffrey Waring | Abbvie, Chicago, IL, United States |
| Fedik Rahimov | Abbvie, Chicago, IL, United States |
| Apinya Lertratanakul | Abbvie, Chicago, IL, United States |
| Nizar Smaoui | Abbvie, Chicago, IL, United States |
| Anne Lehtonen | Abbvie, Chicago, IL, United States |
| David Close | Astra Zeneca, Cambridge, United Kingdom |
| Marla Hochfeld | Bristol Myers Squibb, New York, NY, United States |
| Natalie Bowers | Genentech, San Francisco, CA, United States |
| Rion Pendergrass | Genentech, San Francisco, CA, United States |
| Jorge Esparza Gordillo | GlaxoSmithKline, Brentford, United Kingdom |
| Kirsi Auro | GlaxoSmithKline, Espoo, Finland |
| Dawn Waterworth | Janssen Research & Development, LLC, Spring House, PA, United States |
| Fabiana Farias | Merck, Kenilworth, NJ, United States |
| Kirsi Kalpala | Pfizer, New York, NY, United States |
| Nan Bing | Pfizer, New York, NY, United States |
| Xinli Hu | Pfizer, New York, NY, United States |
| Tarja Laitinen | Pirkanmaa Hospital District, Tampere, Finland |
| Margit Pelkonen | Northern Savo Hospital District, Kuopio, Finland |
| Paula Kauppi | Hospital District of Helsinki and Uusimaa, Helsinki, Finland |
| Hannu Kankaanranta | University of Gothenburg, Gothenburg, Sweden/ Seinäjoki Central Hospital, Seinäjoki, Finland/ Tampere University, Tampere, Finland |
| Terttu Harju | Northern Ostrobothnia Hospital District, Oulu, Finland |
| Riitta Lahesmaa | Hospital District of Southwest Finland, Turku, Finland |
| Nizar Smaoui | Abbvie, Chicago, IL, United States |
| Alex Mackay | Astra Zeneca, Cambridge, United Kingdom |
| Glenda Lassi | Astra Zeneca, Cambridge, United Kingdom |
| Susan Eaton | Biogen, Cambridge, MA, United States |
| Hubert Chen | Genentech, San Francisco, CA, United States |
| Rion Pendergrass | Genentech, San Francisco, CA, United States |
| Natalie Bowers | Genentech, San Francisco, CA, United States |
| Joanna Betts | GlaxoSmithKline, Brentford, United Kingdom |
| Kirsi Auro | GlaxoSmithKline, Espoo, Finland |
| Rajashree Mishra | GlaxoSmithKline, Brentford, United Kingdom |
| Majd Mouded | Novartis, Basel, Switzerland |
| Debby Ngo | Novartis, Basel, Switzerland |
| Teemu Niiranen | Finnish Institute for Health and Welfare (THL), Helsinki, Finland |
| Felix Vaura | Finnish Institute for Health and Welfare (THL), Helsinki, Finland |
| Veikko Salomaa | Finnish Institute for Health and Welfare (THL), Helsinki, Finland |
| Kaj Metsärinne | Hospital District of Southwest Finland, Turku, Finland |
| Jenni Aittokallio | Hospital District of Southwest Finland, Turku, Finland |
| Mika Kähönen | Pirkanmaa Hospital District, Tampere, Finland |
| Jussi Hernesniemi | Pirkanmaa Hospital District, Tampere, Finland |
| Daniel Gordin | Hospital District of Helsinki and Uusimaa, Helsinki, Finland |
| Juha Sinisalo | Hospital District of Helsinki and Uusimaa, Helsinki, Finland |
| Marja-Riitta Taskinen | Hospital District of Helsinki and Uusimaa, Helsinki, Finland |
| Tiinamaija Tuomi | Hospital District of Helsinki and Uusimaa, Helsinki, Finland |
| Timo Hiltunen | Hospital District of Helsinki and Uusimaa, Helsinki, Finland |
| Jari Laukkanen | Central Finland Health Care District, Jyväskylä, Finland |
| Amanda Elliott | Institute for Molecular Medicine Finland (FIMM), HiLIFE, University of Helsinki, Helsinki, Finland; Broad Institute, Cambridge, MA, USA and Massachusetts General Hospital, Boston, MA, USA |
| Mary Pat Reeve | Institute for Molecular Medicine Finland (FIMM), HiLIFE, University of Helsinki, Helsinki, Finland |
| Sanni Ruotsalainen | Institute for Molecular Medicine Finland (FIMM), HiLIFE, University of Helsinki, Helsinki, Finland |
| Benjamin Challis | Astra Zeneca, Cambridge, United Kingdom |
| Dirk Paul | Astra Zeneca, Cambridge, United Kingdom |

| | |
|---|---|
| Julie Hunkapiller | Genentech, San Francisco, CA, United States |
| Natalie Bowers | Genentech, San Francisco, CA, United States |
| Rion Pendergrass | Genentech, San Francisco, CA, United States |
| Audrey Chu | GlaxoSmithKline, Brentford, United Kingdom |
| Kirsi Auro | GlaxoSmithKline, Espoo, Finland |
| Dermot Reilly | Janssen Research & Development, LLC, Boston, MA, United States |
| Mike Mendelson | Novartis, Boston, MA, United States |
| Jaakko Parkkinen | Pfizer, New York, NY, United States |
| Melissa Miller | Pfizer, New York, NY, United States |
| Tuomo Meretoja | Hospital District of Helsinki and Uusimaa, Helsinki, Finland |
| Heikki Joensuu | Hospital District of Helsinki and Uusimaa, Helsinki, Finland |
| Olli Carpén | Hospital District of Helsinki and Uusimaa, Helsinki, Finland |
| Johanna Mattson | Hospital District of Helsinki and Uusimaa, Helsinki, Finland |
| Eveliina Salminen | Hospital District of Helsinki and Uusimaa, Helsinki, Finland |
| Annika Auranen | Pirkanmaa Hospital District , Tampere, Finland |
| Peeter Karihtala | Northern Ostrobothnia Hospital District, Oulu, Finland |
| Päivi Auvinen | Northern Savo Hospital District, Kuopio, Finland |
| Klaus Elenius | Hospital District of Southwest Finland, Turku, Finland |
| Johanna Schleutker | Hospital District of Southwest Finland, Turku, Finland |
| Esa Pitkänen | Institute for Molecular Medicine Finland (FIMM), HiLIFE, University of Helsinki, Helsinki, Finland |
| Nina Mars | Institute for Molecular Medicine Finland (FIMM), HiLIFE, University of Helsinki, Helsinki, Finland |
| Mark Daly | Institute for Molecular Medicine Finland (FIMM), HiLIFE, University of Helsinki, Helsinki, Finland; Broad Institute of MIT and Harvard; Massachusetts General Hospital |
| Relja Popovic | Abbvie, Chicago, IL, United States |
| Jeffrey Waring | Abbvie, Chicago, IL, United States |
| Bridget Riley-Gillis | Abbvie, Chicago, IL, United States |
| Anne Lehtonen | Abbvie, Chicago, IL, United States |
| Jennifer Schutzman | Genentech, San Francisco, CA, United States |
| Julie Hunkapiller | Genentech, San Francisco, CA, United States |
| Natalie Bowers | Genentech, San Francisco, CA, United States |
| Rion Pendergrass | Genentech, San Francisco, CA, United States |
| Diptee Kulkarni | GlaxoSmithKline, Brentford, United Kingdom |
| Kirsi Auro | GlaxoSmithKline, Espoo, Finland |
| Alessandro Porello | Janssen Research & Development, LLC, Spring House, PA, United States |
| Andrey Loboda | Merck, Kenilworth, NJ, United States |
| Heli Lehtonen | Pfizer, New York, NY, United States |
| Stefan McDonough | Pfizer, New York, NY, United States |
| Sauli Vuoti | Janssen-Cilag Oy, Espoo, Finland |
| Kai Kaarniranta | Northern Savo Hospital District, Kuopio, Finland |
| Joni A Turunen | Helsinki University Hospital and University of Helsinki, Helsinki, Finland; Eye Genetics Group, Folkhälsan Research Center, Helsinki, Finland |
| Terhi Ollila | Hospital District of Helsinki and Uusimaa, Helsinki, Finland |
| Hannu Uusitalo | Pirkanmaa Hospital District, Tampere, Finland |
| Juha Karjalainen | Institute for Molecular Medicine Finland (FIMM), HiLIFE, University of Helsinki, Helsinki, Finland |
| Esa Pitkänen | Institute for Molecular Medicine Finland (FIMM), HiLIFE, University of Helsinki, Helsinki, Finland |
| Mengzhen Liu | Abbvie, Chicago, IL, United States |
| Heiko Runz | Biogen, Cambridge, MA, United States |
| Stephanie Loomis | Biogen, Cambridge, MA, United States |
| Erich Strauss | Genentech, San Francisco, CA, United States |
| Natalie Bowers | Genentech, San Francisco, CA, United States |
| Hao Chen | Genentech, San Francisco, CA, United States |
| Rion Pendergrass | Genentech, San Francisco, CA, United States |
| Kaisa Tasanen | Northern Ostrobothnia Hospital District, Oulu, Finland |
| Laura Huilaja | Northern Ostrobothnia Hospital District, Oulu, Finland |
| Katariina Hannula-Jouppi | Hospital District of Helsinki and Uusimaa, Helsinki, Finland |
| Teea Salmi | Pirkanmaa Hospital District, Tampere, Finland |
| Sirkku Peltonen | Hospital District of Southwest Finland, Turku, Finland |
| Leena Koulu | Hospital District of Southwest Finland, Turku, Finland |
| Nizar Smaoui | Abbvie, Chicago, IL, United States |
| Fedik Rahimov | Abbvie, Chicago, IL, United States |
| Anne Lehtonen | Abbvie, Chicago, IL, United States |
| David Choy | Genentech, San Francisco, CA, United States |
| Rion Pendergrass | Genentech, San Francisco, CA, United States |
| Dawn Waterworth | Janssen Research & Development, LLC, Spring House, PA, United States |
| Kirsi Kalpala | Pfizer, New York, NY, United States |
| Ying Wu | Pfizer, New York, NY, United States |
| Pirkko Pussinen | Hospital District of Helsinki and Uusimaa, Helsinki, Finland |
| Aino Salminen | Hospital District of Helsinki and Uusimaa, Helsinki, Finland |
| Tuula Salo | Hospital District of Helsinki and Uusimaa, Helsinki, Finland |
| David Rice | Hospital District of Helsinki and Uusimaa, Helsinki, Finland |
| Pekka Nieminen | Hospital District of Helsinki and Uusimaa, Helsinki, Finland |
| Ulla Palotie | Hospital District of Helsinki and Uusimaa, Helsinki, Finland |
| Maria Siponen | Northern Savo Hospital District, Kuopio, Finland |
| Liisa Suominen | Northern Savo Hospital District, Kuopio, Finland |

| | |
|---|---|
| Päivi Mäntylä | Northern Savo Hospital District, Kuopio, Finland |
| Ulvi Gursoy | Hospital District of Southwest Finland, Turku, Finland |
| Vuokko Anttonen | Northern Ostrobothnia Hospital District, Oulu, Finland |
| Kirsi Sipilä | Research Unit of Oral Health Sciences Faculty of Medicine, University of Oulu, Oulu, Finland; Medical Research Center, Oulu, Oulu University Hospital and University of Oulu, Oulu, Finland |
| Rion Pendergrass | Genentech, San Francisco, CA, United States |
| Hannele Laivuori | Institute for Molecular Medicine Finland (FIMM), HiLIFE, University of Helsinki, Helsinki, Finland |
| Venla Kurra | Pirkanmaa Hospital District, Tampere, Finland |
| Laura Kotaniemi-Talonen | Pirkanmaa Hospital District, Tampere, Finland |
| Oskari Heikinheimo | Hospital District of Helsinki and Uusimaa, Helsinki, Finland |
| Ilkka Kalliala | Hospital District of Helsinki and Uusimaa, Helsinki, Finland |
| Lauri Aaltonen | Hospital District of Helsinki and Uusimaa, Helsinki, Finland |
| Varpu Jokimaa | Hospital District of Southwest Finland, Turku, Finland |
| Johannes Kettunen | Northern Ostrobothnia Hospital District, Oulu, Finland |
| Marja Vääräsmäki | Northern Ostrobothnia Hospital District, Oulu, Finland |
| Outi Uimari | Northern Ostrobothnia Hospital District, Oulu, Finland |
| Laure Morin-Papunen | Northern Ostrobothnia Hospital District, Oulu, Finland |
| Maarit Niinimäki | Northern Ostrobothnia Hospital District, Oulu, Finland |
| Terhi Piltonen | Northern Ostrobothnia Hospital District, Oulu, Finland |
| Katja Kivinen | Institute for Molecular Medicine Finland (FIMM), HiLIFE, University of Helsinki, Helsinki, Finland |
| Elisabeth Widen | Institute for Molecular Medicine Finland (FIMM), HiLIFE, University of Helsinki, Helsinki, Finland |
| Taru Tukiainen | Institute for Molecular Medicine Finland (FIMM), HiLIFE, University of Helsinki, Helsinki, Finland |
| Mary Pat Reeve | Institute for Molecular Medicine Finland (FIMM), HiLIFE, University of Helsinki, Helsinki, Finland |
| Mark Daly | Institute for Molecular Medicine Finland (FIMM), HiLIFE, University of Helsinki, Helsinki, Finland; Broad Institute of MIT and Harvard; Massachusetts General Hospital |
| Niko Välimäki | University of Helsinki, Helsinki, Finland |
| Eija Laakkonen | University of Jyväskylä, Jyväskylä, Finland |
| Jaakko Tyrmi | University of Oulu, Oulu, Finland / University of Tampere, Tampere, Finland |
| Heidi Silven | University of Oulu, Oulu, Finland |
| Eeva Sliz | University of Oulu, Oulu, Finland |
| Riikka Arffman | University of Oulu, Oulu, Finland |
| Susanna Savukoski | University of Oulu, Oulu, Finland |
| Triin Laisk | Estonian biobank, Tartu, Estonia |
| Natalia Pujol | Estonian biobank, Tartu, Estonia |
| Mengzhen Liu | Abbvie, Chicago, IL, United States |
| Bridget Riley-Gillis | Abbvie, Chicago, IL, United States |
| Rion Pendergrass | Genentech, San Francisco, CA, United States |
| Janet Kumar | GlaxoSmithKline, Collegeville, PA, United States |
| Kirsi Auro | GlaxoSmithKline, Espoo, Finland |
| Iiris Hovatta | University of Helsinki, Finland |
| Chia-Yen Chen | Biogen, Cambridge, MA, United States |
| Erkki Isometsä | Hospital District of Helsinki and Uusimaa, Helsinki, Finland |
| Kumar Veerapen | Broad Institute, Cambridge, MA, United States |
| Hanna Ollila | Institute for Molecular Medicine Finland (FIMM), HiLIFE, University of Helsinki, Helsinki, Finland |
| Jaana Suvisaari | Finnish Institute for Health and Welfare (THL), Helsinki, Finland |
| Thomas Damm Als | Aarhus University, Denmark |
| Antti Mäkitie | Department of Otorhinolaryngology - Head and Neck Surgery, University of Helsinki and Helsinki University Hospital, Helsinki, Finland |
| Argyro Bizaki-Vallaskangas | Pirkanmaa Hospital District, Tampere, Finland |
| Sanna Toppila-Salmi | University of Helsinki, Finland |
| Tytti Willberg | Hospital District of Southwest Finland, Turku, Finland |
| Elmo Saarentaus | Institute for Molecular Medicine Finland (FIMM), HiLIFE, University of Helsinki, Helsinki, Finland |
| Antti Aarnisalo | Hospital District of Helsinki and Uusimaa, Helsinki, Finland |
| Eveliina Salminen | Hospital District of Helsinki and Uusimaa, Helsinki, Finland |
| Elisa Rahikkala | Northern Ostrobothnia Hospital District, Oulu, Finland |
| Johannes Kettunen | Northern Ostrobothnia Hospital District, Oulu, Finland |
| Kristiina Aittomäki | Department of Medical Genetics, Helsinki University Central Hospital, Helsinki, Finland |
| Fredrik Åberg | Transplantation and Liver Surgery Clinic, Helsinki University Hospital, Helsinki University, Helsinki, Finland |
| Mitja Kurki | Institute for Molecular Medicine Finland (FIMM), HiLIFE, University of Helsinki, Helsinki, Finland; Broad Institute, Cambridge, MA, United States |
| Samuli Ripatti | Institute for Molecular Medicine Finland (FIMM), HiLIFE, University of Helsinki, Helsinki, Finland |
| Mark Daly | Institute for Molecular Medicine, Finland (FIMM), HiLIFE, University of Helsinki, Helsinki, Finland; Broad Institute of MIT and Harvard; Massachusetts General Hospital |
| Juha Karjalainen | Institute for Molecular Medicine Finland (FIMM), HiLIFE, University of Helsinki, Helsinki, Finland |
| Aki Havulinna | Institute for Molecular Medicine Finland (FIMM), HiLIFE, University of Helsinki, Helsinki, Finland; Finnish Institute for Health and Welfare (THL), Helsinki, Finland |
| Juha Mehtonen | Institute for Molecular Medicine Finland (FIMM), HiLIFE, University of Helsinki, Helsinki, Finland |
| Priit Palta | Institute for Molecular Medicine Finland (FIMM), HiLIFE, University of Helsinki, Helsinki, Finland |
| Shabbeer Hassan | Institute for Molecular Medicine Finland (FIMM), HiLIFE, University of Helsinki, Helsinki, Finland |

| | |
|---|---|
| Pietro Della Briotta Parolo | Institute for Molecular Medicine Finland (FIMM), HiLIFE, University of Helsinki, Helsinki, Finland |
| Wei Zhou | Broad Institute, Cambridge, MA, United States |
| Mutaamba Maasha | Broad Institute, Cambridge, MA, United States |
| Kumar Veerapen | Broad Institute, Cambridge, MA, United States |
| Shabbeer Hassan | Institute for Molecular Medicine Finland (FIMM), HiLIFE, University of Helsinki, Helsinki, Finland |
| Susanna Lemmelä | Institute for Molecular Medicine Finland (FIMM), HiLIFE, University of Helsinki, Helsinki, Finland |
| Manuel Rivas | University of Stanford, Stanford, CA, United States |
| Mari E. Niemi | Institute for Molecular Medicine Finland (FIMM), HiLIFE, University of Helsinki, Helsinki, Finland |
| Aarno Palotie | Institute for Molecular Medicine Finland (FIMM), HiLIFE, University of Helsinki, Helsinki, Finland |
| Aoxing Liu | Institute for Molecular Medicine Finland (FIMM), HiLIFE, University of Helsinki, Helsinki, Finland |
| Arto Lehisto | Institute for Molecular Medicine Finland (FIMM), HiLIFE, University of Helsinki, Helsinki, Finland |
| Andrea Ganna | Institute for Molecular Medicine Finland (FIMM), HiLIFE, University of Helsinki, Helsinki, Finland |
| Vincent Llorens | Institute for Molecular Medicine Finland (FIMM), HiLIFE, University of Helsinki, Helsinki, Finland |
| Hannele Laivuori | Institute for Molecular Medicine Finland (FIMM), HiLIFE, University of Helsinki, Helsinki, Finland |
| Taru Tukiainen | Institute for Molecular Medicine Finland (FIMM), HiLIFE, University of Helsinki, Helsinki, Finland |
| Mary Pat Reeve | Institute for Molecular Medicine Finland (FIMM), HiLIFE, University of Helsinki, Helsinki, Finland |
| Henrike Heyne | Institute for Molecular Medicine Finland (FIMM), HiLIFE, University of Helsinki, Helsinki, Finland |
| Nina Mars | Institute for Molecular Medicine Finland (FIMM), HiLIFE, University of Helsinki, Helsinki, Finland |
| Joel Rämö | Institute for Molecular Medicine Finland (FIMM), HiLIFE, University of Helsinki, Helsinki, Finland |
| Elmo Saarentaus | Institute for Molecular Medicine Finland (FIMM), HiLIFE, University of Helsinki, Helsinki, Finland |
| Hanna Ollila | Institute for Molecular Medicine Finland (FIMM), HiLIFE, University of Helsinki, Helsinki, Finland |
| Rodos Rodosthenous | Institute for Molecular Medicine Finland (FIMM), HiLIFE, University of Helsinki, Helsinki, Finland |
| Satu Strausz | Institute for Molecular Medicine Finland (FIMM), HiLIFE, University of Helsinki, Helsinki, Finland |
| Tuula Palotie | University of Helsinki and Hospital District of Helsinki and Uusimaa, Helsinki, Finland |
| Kimmo Palin | University of Helsinki, Helsinki, Finland |
| Javier Garcia-Tabuenca | University of Tampere, Tampere, Finland |
| Harri Siirtola | University of Tampere, Tampere, Finland |
| Tuomo Kiiskinen | Institute for Molecular Medicine Finland (FIMM), HiLIFE, University of Helsinki, Helsinki, Finland |
| Jiwoo Lee | Institute for Molecular Medicine Finland (FIMM), HiLIFE, University of Helsinki, Helsinki, Finland; Broad Institute, Cambridge, MA, United States |
| Kristin Tsuo | Institute for Molecular Medicine Finland (FIMM), HiLIFE, University of Helsinki, Helsinki, Finland; Broad Institute, Cambridge, MA, United States |
| Amanda Elliott | Institute for Molecular Medicine Finland (FIMM), HiLIFE, University of Helsinki, Helsinki, Finland; Broad Institute, Cambridge, MA, USA and Massachusetts General Hospital, Boston, MA, USA |
| Kati Kristiansson | THL Biobank / Finnish Institute for Health and Welfare (THL), Helsinki, Finland |
| Mikko Arvas | Finnish Red Cross Blood Service / Finnish Hematology Registry and Clinical Biobank, Helsinki, Finland |
| Kati Hyvärinen | Finnish Red Cross Blood Service, Helsinki, Finland |
| Jarmo Ritari | Finnish Red Cross Blood Service, Helsinki, Finland |
| Olli Carpén | Helsinki Biobank / Helsinki University and Hospital District of Helsinki and Uusimaa, Helsinki |
| Johannes Kettunen | Northern Finland Biobank Borealis / University of Oulu / Northern Ostrobothnia Hospital District, Oulu, Finland |
| Katri Pylkäs | University of Oulu, Oulu, Finland |
| Eeva Sliz | University of Oulu, Oulu, Finland |
| Minna Karjalainen | University of Oulu, Oulu, Finland |
| Tuomo Mantere | Northern Finland Biobank Borealis / University of Oulu / Northern Ostrobothnia Hospital District, Oulu, Finland |
| Eeva Kangasniemi | Finnish Clinical Biobank Tampere / University of Tampere / Pirkanmaa Hospital District, Tampere, Finland |
| Sami Heikkinen | University of Eastern Finland, Kuopio, Finland |
| Arto Mannermaa | Biobank of Eastern Finland / University of Eastern Finland / Northern Savo Hospital District, Kuopio, Finland |
| Eija Laakkonen | University of Jyväskylä, Jyväskylä, Finland |
| Nina Pitkänen | Auria Biobank / University of Turku / Hospital District of Southwest Finland, Turku, Finland |
| Samuel Lessard | Translational Sciences, Sanofi R&D, Framingham, MA, USA |
| Clément Chatelain | Translational Sciences, Sanofi R&D, Framingham, MA, USA |
| Perttu Terho | Auria Biobank / University of Turku / Hospital District of Southwest Finland, Turku, Finland |
| Sirpa Soini | THL Biobank / Finnish Institute for Health and Welfare (THL), Helsinki, Finland |
| Jukka Partanen | Finnish Red Cross Blood Service / Finnish Hematology Registry and Clinical Biobank, Helsinki, Finland |
| Eero Punkka | Helsinki Biobank / Helsinki University and Hospital District of Helsinki and Uusimaa, Helsinki |
| Raisa Serpi | Northern Finland Biobank Borealis / University of Oulu / Northern Ostrobothnia Hospital District, Oulu, Finland |
| Sanna Siltanen | Finnish Clinical Biobank Tampere / University of Tampere / Pirkanmaa Hospital District, Tampere, Finland |
| Veli-Matti Kosma | Biobank of Eastern Finland / University of Eastern Finland / Northern Savo Hospital District, Kuopio, Finland |
| Teijo Kuopio | Central Finland Biobank / University of Jyväskylä / Central Finland Health Care District, Jyväskylä, Finland |
| Anu Jalanko | Institute for Molecular Medicine Finland (FIMM), HiLIFE, University of Helsinki, Helsinki, Finland |
| Huei-Yi Shen | Institute for Molecular Medicine Finland (FIMM), HiLIFE, University of Helsinki, Helsinki, Finland |
| Risto Kajanne | Institute for Molecular Medicine Finland (FIMM), HiLIFE, University of Helsinki, Helsinki, Finland |

| | |
|---|---|
| Mervi Aavikko | Institute for Molecular Medicine Finland (FIMM), HiLIFE, University of Helsinki, Helsinki, Finland |
| Mitja Kurki | Institute for Molecular Medicine Finland (FIMM), HiLIFE, University of Helsinki, Helsinki, Finland; Broad Institute, Cambridge, MA, United States |
| Juha Karjalainen | Institute for Molecular Medicine Finland (FIMM), HiLIFE, University of Helsinki, Helsinki, Finland |
| Pietro Della Briotta Parolo | Institute for Molecular Medicine Finland (FIMM), HiLIFE, University of Helsinki, Helsinki, Finland |
| Arto Lehisto | Institute for Molecular Medicine Finland (FIMM), HiLIFE, University of Helsinki, Helsinki, Finland |
| Juha Mehtonen | Institute for Molecular Medicine Finland (FIMM), HiLIFE, University of Helsinki, Helsinki, Finland |
| Wei Zhou | Broad Institute, Cambridge, MA, United States |
| Masahiro Kanai | Broad Institute, Cambridge, MA, United States |
| Mutaamba Maasha | Broad Institute, Cambridge, MA, United States |
| Kumar Veerapen | Broad Institute, Cambridge, MA, United States |
| Hannele Laivuori | Institute for Molecular Medicine Finland (FIMM), HiLIFE, University of Helsinki, Helsinki, Finland |
| Aki Havulinna | Institute for Molecular Medicine Finland (FIMM), HiLIFE, University of Helsinki, Helsinki, Finland; Finnish Institute for Health and Welfare (THL), Helsinki, Finland |
| Susanna Lemmelä | Institute for Molecular Medicine Finland (FIMM), HiLIFE, University of Helsinki, Helsinki, Finland |
| Tuomo Kiiskinen | Institute for Molecular Medicine Finland (FIMM), HiLIFE, University of Helsinki, Helsinki, Finland |
| L. Elisa Lahtela | Institute for Molecular Medicine Finland (FIMM), HiLIFE, University of Helsinki, Helsinki, Finland |
| Mari Kaunisto | Institute for Molecular Medicine Finland (FIMM), HiLIFE, University of Helsinki, Helsinki, Finland |
| Elina Kilpeläinen | Institute for Molecular Medicine Finland (FIMM), HiLIFE, University of Helsinki, Helsinki, Finland |
| Timo P. Sipilä | Institute for Molecular Medicine Finland (FIMM), HiLIFE, University of Helsinki, Helsinki, Finland |
| Oluwaseun Alexander Dada | Institute for Molecular Medicine Finland (FIMM), HiLIFE, University of Helsinki, Helsinki, Finland |
| Awaisa Ghazal | Institute for Molecular Medicine Finland (FIMM), HiLIFE, University of Helsinki, Helsinki, Finland |
| Anastasia Kytölä | Institute for Molecular Medicine Finland (FIMM), HiLIFE, University of Helsinki, Helsinki, Finland |
| Rigbe Weldatsadik | Institute for Molecular Medicine Finland (FIMM), HiLIFE, University of Helsinki, Helsinki, Finland |
| Kati Donner | Institute for Molecular Medicine Finland (FIMM), HiLIFE, University of Helsinki, Helsinki, Finland |
| Timo P. Sipilä | Institute for Molecular Medicine Finland (FIMM), HiLIFE, University of Helsinki, Helsinki, Finland |
| Anu Loukola | Helsinki Biobank / Helsinki University and Hospital District of Helsinki and Uusimaa, Helsinki |
| Päivi Laiho | THL Biobank / Finnish Institute for Health and Welfare (THL), Helsinki, Finland |
| Tuuli Sistonen | THL Biobank / Finnish Institute for Health and Welfare (THL), Helsinki, Finland |
| Essi Kaiharju | THL Biobank / Finnish Institute for Health and Welfare (THL), Helsinki, Finland |
| Markku Laukkanen | THL Biobank / Finnish Institute for Health and Welfare (THL), Helsinki, Finland |
| Elina Järvensivu | THL Biobank / Finnish Institute for Health and Welfare (THL), Helsinki, Finland |
| Sini Lähteenmäki | THL Biobank / Finnish Institute for Health and Welfare (THL), Helsinki, Finland |
| Lotta Männikkö | THL Biobank / Finnish Institute for Health and Welfare (THL), Helsinki, Finland |
| Regis Wong | THL Biobank / Finnish Institute for Health and Welfare (THL), Helsinki, Finland |
| Auli Toivola | THL Biobank / Finnish Institute for Health and Welfare (THL), Helsinki, Finland |
| Minna Brunfeldt | THL Biobank / Finnish Institute for Health and Welfare (THL), Helsinki, Finland |
| Hannele Mattsson | THL Biobank / Finnish Institute for Health and Welfare (THL), Helsinki, Finland |
| Kati Kristiansson | THL Biobank / Finnish Institute for Health and Welfare (THL), Helsinki, Finland |
| Susanna Lemmelä | Institute for Molecular Medicine Finland (FIMM), HiLIFE, University of Helsinki, Helsinki, Finland |
| Sami Koskelainen | THL Biobank / Finnish Institute for Health and Welfare (THL), Helsinki, Finland |
| Tero Hiekkalinna | THL Biobank / Finnish Institute for Health and Welfare (THL), Helsinki, Finland |
| Teemu Paajanen | THL Biobank / Finnish Institute for Health and Welfare (THL), Helsinki, Finland |
| Priit Palta | Institute for Molecular Medicine Finland (FIMM), HiLIFE, University of Helsinki, Helsinki, Finland |
| Kalle Pärn | Institute for Molecular Medicine Finland (FIMM), HiLIFE, University of Helsinki, Helsinki, Finland |
| Mart Kals | Institute for Molecular Medicine Finland (FIMM), HiLIFE, University of Helsinki, Helsinki, Finland |
| Shuang Luo | Institute for Molecular Medicine Finland (FIMM), HiLIFE, University of Helsinki, Helsinki, Finland |
| Vishal Sinha | Institute for Molecular Medicine Finland (FIMM), HiLIFE, University of Helsinki, Helsinki, Finland |
| Tarja Laitinen | Pirkanmaa Hospital District, Tampere, Finland |
| Mary Pat Reeve | Institute for Molecular Medicine Finland (FIMM), HiLIFE, University of Helsinki, Helsinki, Finland |
| Marianna Niemi | University of Tampere, Tampere, Finland |
| Kumar Veerapen | Broad Institute, Cambridge, MA, United States |
| Harri Siirtola | University of Tampere, Tampere, Finland |
| Javier Gracia-Tabuenca | University of Tampere, Tampere, Finland |
| Mika Helminen | University of Tampere, Tampere, Finland |
| Tiina Luukkaala | University of Tampere, Tampere, Finland |
| Iida Vähätalo | University of Tampere, Tampere, Finland |
| Jyrki Pitkänen | Institute for Molecular Medicine Finland (FIMM), HiLIFE, University of Helsinki, Helsinki, Finland |
| Marco Hautalahti | Finnish Biobank Cooperative - FINBB |
| Johanna Mäkelä | Finnish Biobank Cooperative - FINBB |
| Sarah Smith | Finnish Biobank Cooperative - FINBB |
| Tom Southerington | Finnish Biobank Cooperative - FINBB |
